# Wastewater and seroprevalence for pandemic preparedness: variant analysis, vaccination effect, and hospitalization forecasting for SARS-CoV-2, in Jefferson County, Kentucky

**DOI:** 10.1101/2023.01.06.23284260

**Authors:** Rochelle H. Holm, Grzegorz A. Rempala, Boseung Choi, J. Michael Brick, Alok R. Amraotkar, Rachel J. Keith, Eric C. Rouchka, Julia H. Chariker, Kenneth E. Palmer, Ted Smith, Aruni Bhatnagar

## Abstract

Despite wide scale assessments, it remains unclear how large-scale SARS-CoV-2 vaccination affected the wastewater concentration of the virus or the overall disease burden as measured by hospitalization rates. We used weekly SARS-CoV-2 wastewater concentration with a stratified random sampling of seroprevalence, and linked vaccination and hospitalization data, from April 2021–August 2021 in Jefferson County, Kentucky (USA). Our susceptible (*S*), vaccinated (*V*), variant-specific infected (*I*_1_ and *I*_2_), recovered (*R*), and seropositive (*T*) model (*SVI*_2_ *RT*) tracked prevalence longitudinally. This was related to wastewater concentration. The 64% county vaccination rate translated into about 61% decrease in SARS-CoV-2 incidence. The estimated effect of SARS-CoV-2 Delta variant emergence was a 24-fold increase of infection counts, which corresponded to an over 9-fold increase in wastewater concentration. Hospitalization burden and wastewater concentration had the strongest correlation (r = 0.95) at 1 week lag. Our study underscores the importance of continued environmental surveillance post-vaccine and provides a proof-of-concept for environmental epidemiology monitoring of infectious disease for future pandemic preparedness.

## 1. Introduction

In the wake of the COVID-19 pandemic, more reliable methods of measuring disease prevalence in communities are urgently needed, particularly methods that do not involve the expensive and cumbersome process of collecting individual level data. Completed development and validation of such methods are likely to be a center piece of preparedness for future pandemics. Wastewater concentration, when properly calibrated, can be a surrogate for estimates based on community prevalence of infection.^1–3^ Moreover, wastewater-based epidemiology offers the opportunity of estimating community disease prevalence even with asymptomatic disease.^2,3^ A handful of previous evaluations of the relationship between SARS-CoV-2 wastewater concentration and the COVIDLJ19 vaccine have relied almost exclusively on statistical models calibrated with case counts or other convenience sampling.^4–8^ These data run the risk of biased underrepresentation of asymptomatic individuals who may not seek testing, or individuals testing in settings where reporting is low or not required.^9^ Other mathematical models are based at a state or national spatiotemporal scale.^10–13^ Hence, in this study we consider this question in the context of randomized seroprevalence surveillance, combining mechanistic and statistical frameworks to obtain more robust and realistic estimates of changes in disease prevalence.

We address the question of how changes in the concentrations during the Alpha and Delta variant waves of the pandemic affected wastewater concentrations by looking in detail at a small geographical area which other studies have not done previously. For our analysis, we used repeated cross-sectional community-wide stratified randomized sampling to measure SARS-CoV-2 nucleocapsid (N1) specific IgG antibody-based seroprevalence in Jefferson County, Kentucky (USA), from April 2021 through August 2021 to determine post-vaccine community prevalence at a sub-county scale. We then related this to a statistical linear model and the available sub-county weekly wastewater surveillance data which yielded estimates of the explicit impact of vaccination and seroimmunity on a SARS-CoV-2 wastewater concentration estimate, while controlling for prevalence in different epidemic phases using a population level ecological model. The latter may be easily translated into other important public health indicators such as the patterns of hospitalization. The ecological model *SVI*_2_ *RT* was used to longitudinally monitor the proportions of individuals in various health stages. These included those who were susceptible (*S*), vaccinated (*V*), infected with non-Delta variant (*I*_1_), infected with Delta variant (*I*_2_), recovered (*R*), or seropositive (*T*).

## 2. Results

### 2.1. Wastewater regression

When examining the relationship between the prevalence estimated from the _2_ model and the observed wastewater levels the results of the Bayesian regression analysis (Supplemental Material Appendix C) demonstrate a significant trend. This analysis considers both countywide aggregation and five localized sewershed locations allowing finer geographic resolution. The trend is effectively summarized by the corresponding posterior regression line. To obtain reliable and stable longitudinal concentration readings, the concentration of SARS-CoV-2 (N1) was normalized by pepper mild mottle virus (PMMoV) concentration to enhance accuracy and precision and minimize variance in assessing changes in the concentration of the virus over time.

To assess the impact of prevalence on observed wastewater concentration for the Alpha and Delta variants, we employed a regression model known as the broken stick regression to account for variant-specific patterns of virus shedding and infection rates. This model incorporates two regressors: one for the Alpha variant with an estimated prevalence prior to 5 June 2021, and another for the Delta variant with an estimated prevalence after 5 June 2021. Consequently, the model encompasses two different regression coefficients corresponding to the respective prevalence. The transition date was determined by the prevailing dominance of the Alpha and Delta variants as inferred from wastewater samples. For the aggregate model, the estimated intercept is -4.222 × 10^−4^ (CI = (−9.458 × 10^−4^, 7.921 × 10^−5^)) and the two slopes are 0.815 (CI = (−0.023, 1.717)) and 0.385 (CI = (0.318, 0.455)) (Table S5). Overall, the regression model fits the data well (*R*^2^ = 0.90).

### 2.2. Effect of vaccination on disease incidence and wastewater concentration

We first compared the estimated incidence of the *SVI*_2_ *RT* model under two different vaccination scenarios (factual 64% vaccination rate and counterfactual 0% vaccination rate) while adjusting for the Delta variant emergence (Figure 1). The peak and the overall temporal dynamics are different under the two scenarios across each location, credible intervals for the incidence with and without vaccination are overlapping and indicate that the scenario curves could have statistically close values during certain times. To quantify these differences more precisely, we computed the location-specific vaccination effects as the ratios of the areas under the two scenario incidence curves. Specifically, we compared the area under the curve (corresponding to a relative cumulative incidence) for the “with-vaccination” scenario to that of the “without-vaccination” scenario. The value obtained for the aggregated data was 0.390, with the remaining sewershed specific effects being even stronger at 0.502, 0.393, and 0.479 for MSD1, MSD2, and MSD3 to 5, respectively. Based on converting these ratios to excess incidence, we estimate that without vaccination, the reported integrated incidence may have increased about 156.2% (CI = (95.2%, 175.7%)) above the observed level in Jefferson County (Figure 1; panel A) and about 99.4% (CI = (94.2%, 108.5%)), 154.5% (CI = (3.2%, 154.7%)), and 108.8% (CI = (52.8%, 109.2%)) in different sewershed areas (Figure 1 panels B–D; Table S7).

**Figure 1.**
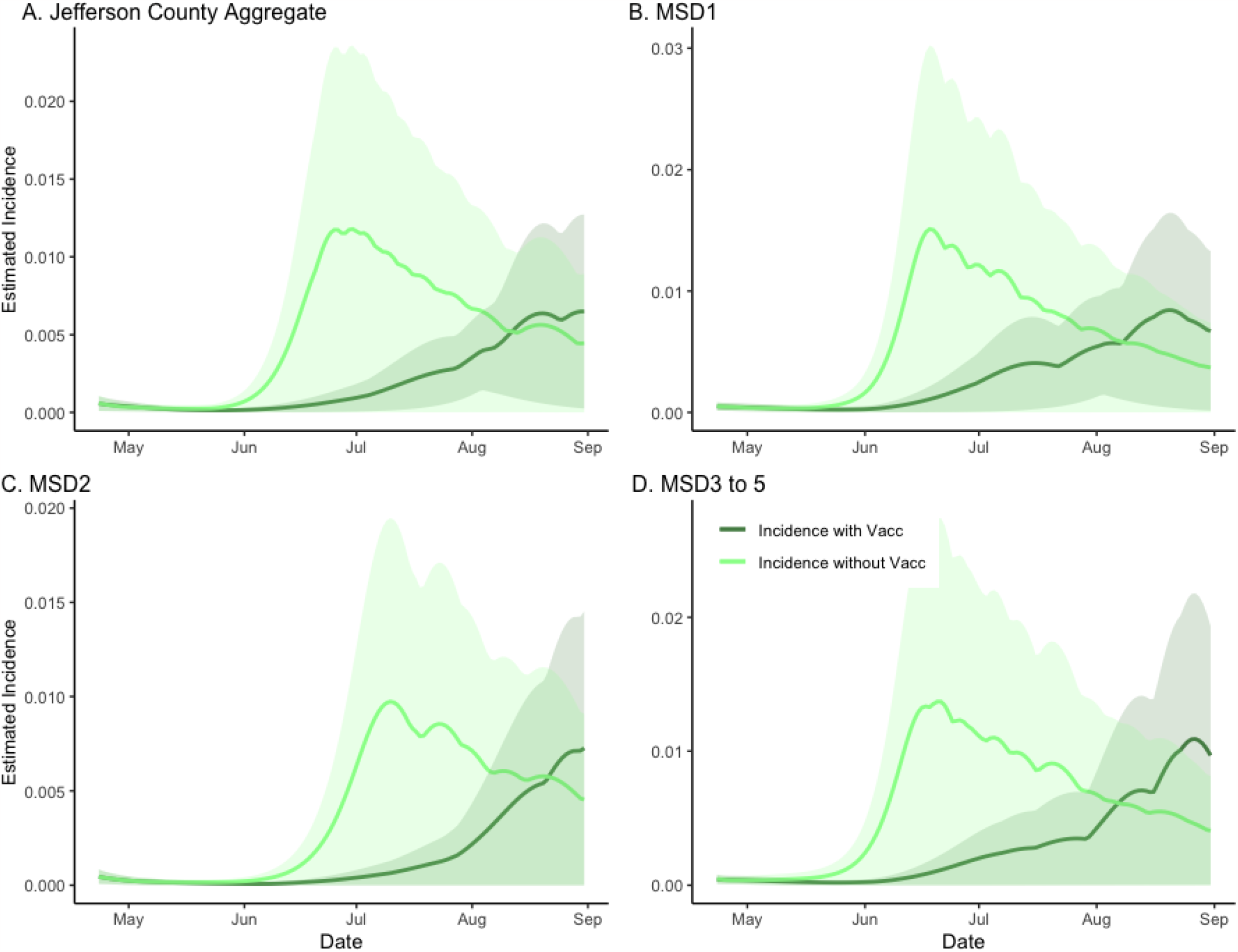
The estimated effect of vaccination on incidence in sewersheds of Jefferson County, KY (USA). The dark green line is the factual *SVI*_2_*RT* model estimated incidence (with vaccination), and the light green line is the corresponding counterfactual estimated incidence with vaccination effect zeroed out. The shaded areas represent 95% credible intervals. The panels compare the vaccination effect in Jefferson County (Panel A) as well as stratified by sewershed (Panels B–D).

To obtain estimates of the effects of vaccination on the wastewater concentration of the virus, we developed a hybrid inferential model combining the wastewater regression equation with the *SVI*_2_*RT* estimated prevalence, under two different vaccination scenarios (factual 64% rate and counterfactual 0% rate) (Figure 2). The use of *SVI*_2_ *RT* (which accounts for the effect of different virulence of the two different SARS-CoV-2 variants) automatically adjusted our analysis for the Delta variant emergence. Because the estimated prevalence from the *SVI*_2_ *RT* model and the normalized wastewater concentration are highly correlated, the hybrid model is seen to fit the data well. As before, to quantify the location-specific vaccination effects, we calculated the location-specific ratios under two curves in an analogous way as when quantifying the vaccination effect on the disease incidence. The ratios of the areas under the two curves, under factual (vaccinated) and counterfactual (unvaccinated) scenarios, were computed. The Jefferson County (Figure 2; panel A) ratio was equal to 0.314, and the remaining sewershed location ratios (Figure 2; panels B–D) were equal to, respectively, 0.448, 0.330, and 0.375. The estimate of excess wastewater virus without vaccination is estimated as 218.9% (CI = (193.5%, 242.4%)), 123.1% (CI = (105.0%, 144.0%)), 202.8% (CI = (192.8%, 203.4%)), and 166.9%, (CI = (146.6%, 187.1%)) respectively (Table S7).

**Figure 2.**
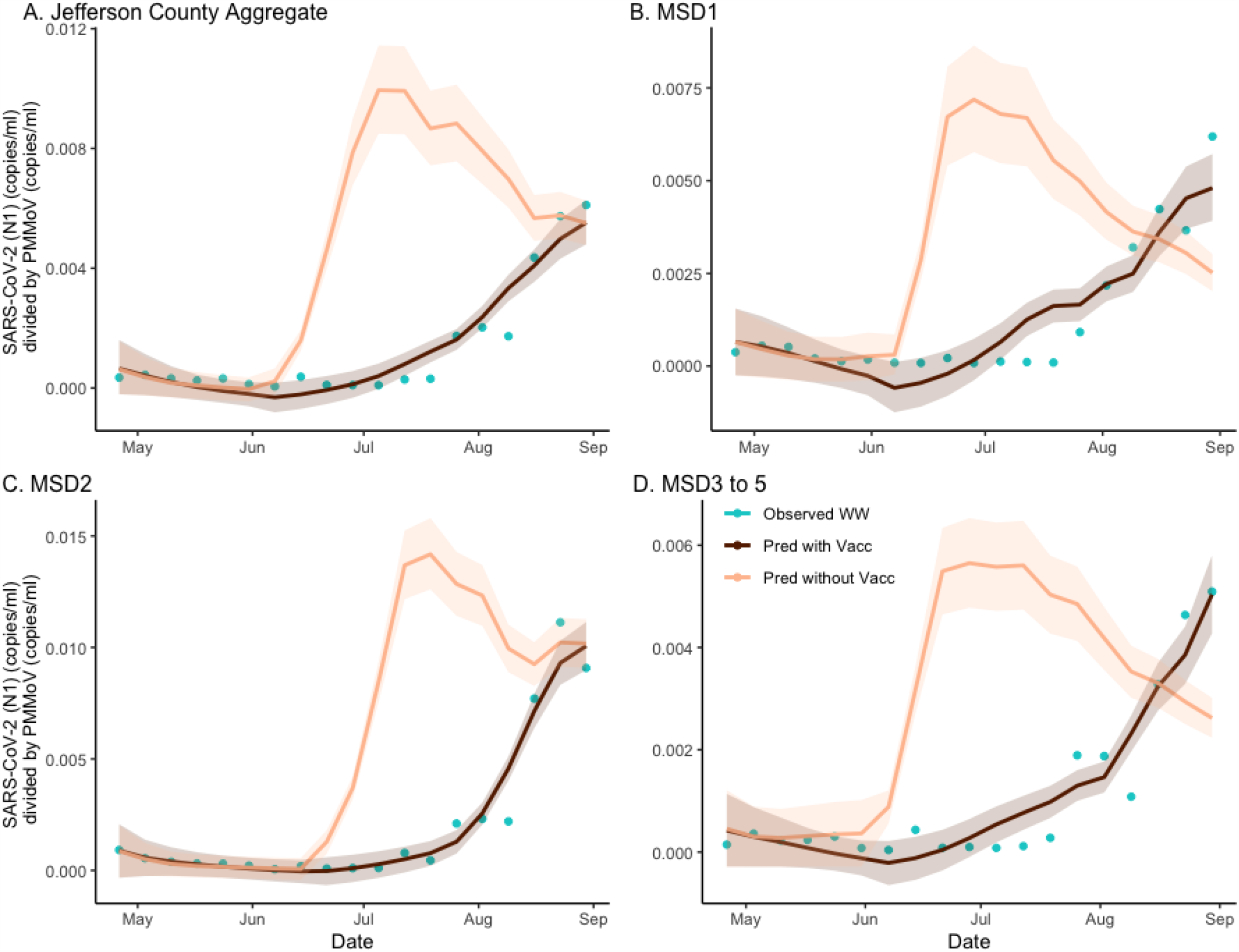
The estimated effect of vaccination on SARS-CoV-2 (N1) wastewater concentration normalized by pepper mild mottle virus in sewersheds of Jefferson County, KY (USA). The dark brown line is the regression-based fit to the wastewater concentration data and the light brown line is the prediction of wastewater concentration using synthetic prevalence from *SVI*_2_ *RT* model with vaccination effect zeroed out. The shaded areas represent 95% credible intervals. The blue dots are observed weekly average wastewater concentrations. The panels compare the vaccination effect on wastewater concentration for Jefferson County (Panel A) as well as stratified by sewershed (Panels B–D).

### 2.3. Effects of virus variant on disease incidence and wastewater concentration

Alpha was the dominant variant at the start of our study period on 30 March 2021. The Delta variant was first introduced into the two largest urban sewersheds as the dominant variant on 12 July 2021 and appearing in the more rural sewersheds in the following two-week period. More recently, we have reported on the re-emergence of Delta in the MSD3 site during the Omicron wave,^14^ which indicates the persistence of specific variants in wastewater can be variable and are likely influenced by several factors, including the rates of incidence and vaccination.

In our analysis, we assumed a 50% higher infectivity of the SARS-CoV-2 Delta variant in comparison with its Alpha predecessor.^15^ In the counterfactual model, where only the Alpha variant was present, the epidemic was seen to dissipate, indicating the effective reproduction number of less than one. This was in contrast with the factual, full *SVI*_2_ *RT* model fit (with both Alpha- and Delta-variants present), where the incidence was seen to rise rapidly. As in the previous section, to quantify the difference between the two curves, which we interpreted as measuring the effect of introducing the Delta variant, we calculated the ratio of areas under the two curves, obtaining the values of 23.524, 31.103, 23.986, and 33.336 for the aggregate, MSD1, MSD2, and MSD3–5 regions respectively (Figure 3; panels A–D). The estimate of the decrease in total incidence without the variant was found as 95.75% (CI = (95.74%, 95.91%)), 96.78% (CI = (95.54%, 96.84%)), 95.8% (CI = (2.7%, 96.0%)), and 97.0% (CI = (38.6%, 97.1%)), respectively. Note that the two lower bounds of the ratio for the MSD2 and MSD3–5 areas were relatively small. This is because the estimated incidence from both variants had lower CI areas that were close to zero.

**Figure 3.**
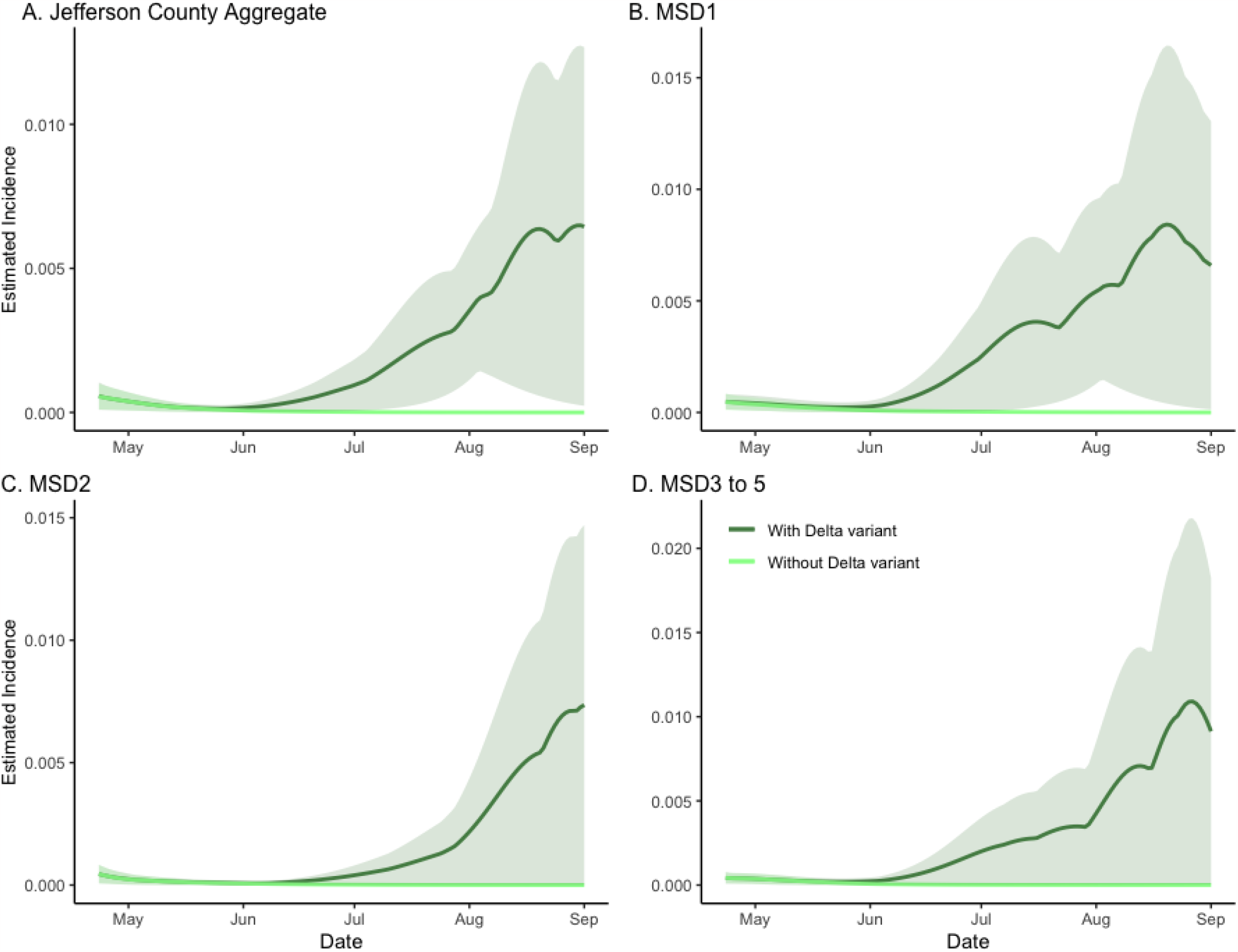
The model-based analysis of the Delta variant effect on SARS-CoV-2 incidence rate estimates in sewersheds of Jefferson County, KY (USA). The dark green line is the estimated factual full model incidence (both Alpha and Delta variants present), and the light green line is the counterfactual incidence estimated from the model with no Delta variant. The shaded areas represent 95% credible intervals. The panels compare the incidence rate in Jefferson County (Panel A) as well as stratified by sewershed (Panels B–D).

To identify the effect of the Delta variant emergence on the observed wastewater concentration, we again applied the hybrid model from the previous section. Genetic variants can have an impact on fecal shedding.^16^ In the current analysis, the regression model was applied to predict the longitudinal wastewater concentrations from both factual (both variants present) and counterfactual prevalence data (no Delta variant) (Figure 4). As with the analysis of the vaccination effects, here we also considered the ratios of areas under the corresponding curves as measures of Delta variant effects in specific locations. Based on the aggregated ratio values of 8.655, and on the location-specific ratio values 5.695, 9.675, and 8.530, the estimate of excess wastewater concentration due to Delta was found as 88.4% (CI = (87.7%, 88.7%)), 82.4% (CI = (81.4%, 84.0%)), 89.7% (CI = (88.5%, 90.8%)), and 88.3% (CI = (87.3%, 89.1%)) respectively (Table S7). By utilizing the fitted regression coefficients (see Section S3.2), we can further examine the impact of the Alpha and Delta variants on wastewater concentrations. To facilitate a comparison, we employed standardized regression coefficients instead of the original scale. Because the range of the Alpha variant prevalence and the number of data points of the Alpha variant are smaller than that of the Delta variant, the slope coefficient of the Alpha variant is larger than that of the Delta variant but is not significant. For the aggregated model (Figure 4; panel A), the standardized regression coefficient of the Alpha variant prevalence is 3.464 × 10^−4^ (CI = (4.460 × 10^−7^, 6.946 × 10^−4^)) and the Delta variant prevalence is 1.992 × 10^−3^ (CI = (1.627 × 10^−3^, 2.344 × 10^−3^)). Hence, the effect of the Delta variant was found to be 5.8 times greater than that of the Alpha variant. The fitted line of wastewater concentration exhibits a transition point, and the broken stick regression line aligns well with the data (R-square value 0.904). We can also see similar patterns in other sewershed locations (Figure 4; panels B–D). The standardized regression coefficients for each sewershed area are 5.053 × 10^−4^ (CI = (1.081 × 10^−5^, 9.796 × 10^−4^)) and 1.880 × 10^−3^ (CI = (1.387 × 10^−3^, 2.339 × 10^−3^)) for MSD1, 3.586 × 10^−4^ (CI = (−1.148 × 10^−4^, 8.337 × 10^−4^)) and 3.395 × 10^−3^ (CI = (2.921 × 10^−3^, 3.872 × 10^−3^)) for MSD2, and 2.518 × 10^−4^ (CI = (−4.963 × 10^−5^, 5.616 × 10^−4^)) and 1.609 × 10^−3^ (CI = (1.291 × 10^−3^, 1.910 × 10^−3^)) for MSD3 to 5 respectively.

**Figure 4.**
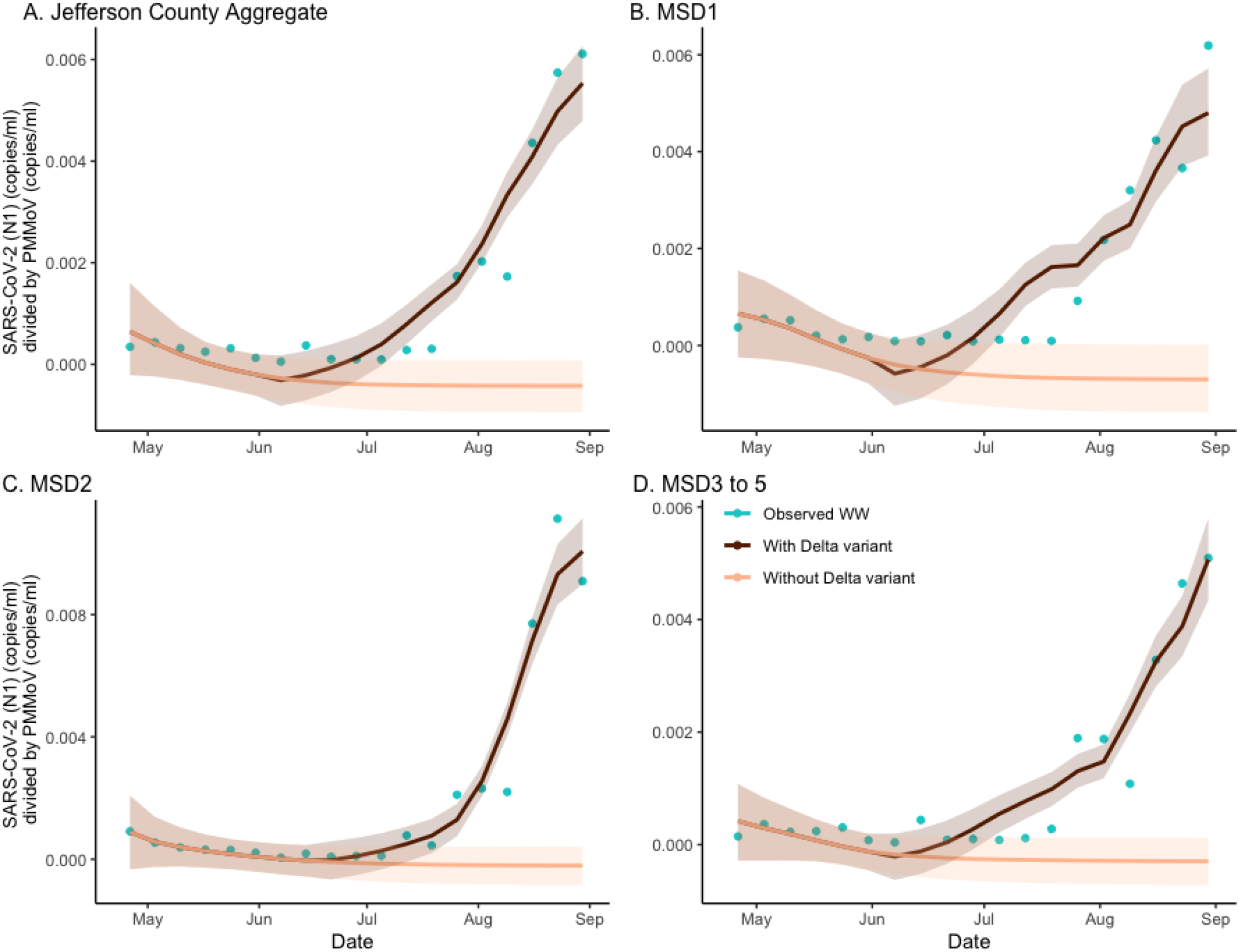
The estimated effect of Alpha and Delta variant on SARS-CoV-2 (N1) wastewater concentration normalized by pepper mild mottle virus in sewersheds of Jefferson County, KY (USA). The dark brown line is the regression-based fit to the wastewater concentration with the Alpha and Delta variant and the light brown line is the prediction of wastewater concentration using synthetic prevalence from the *SVI*_2_ *RT* model with the Alpha variant only. The shaded areas represent 95% credible intervals. The blue dots are observed weekly average wastewater concentration. The panels compare the variant effect on wastewater concentration for Jefferson County (Panel A) as well as stratified by sewershed (Panels B–D).

### 2.4. Forecasting hospitalization rates based on wastewater concentration

Hospitalization estimates under both vaccinated (64% vaccination rate^17^) and unvaccinated (0% vaccination rate) scenarios were obtained by applying a hierarchical regression model where we first regressed wastewater concentration on the *SVI*_2_ *RT* model prevalence and then regressed hospitalization counts on the wastewater concentrations (Figure 5). As hospitalization is likely to occur sometime after symptom onset, we considered a range of no lag to 5-week lag period. A 1-week lagged-regression model was the best fit where the length of the lag time was based on the overall model fit criteria. The fitted intercept and slope coefficients were 1.222 × 10^−4^ (std = 3.345 × 10^−5^) and 0.181 (std = 0.0150) for vaccinated and unvaccinated scenarios respectively (R-square of 0.895) (Figure 5; panel A). The maximal number of the observed daily average hospitalizations under vaccination scenario was 110.4 per weekly average (actual 122.0 in daily) at the end of August. However, without vaccination, the maximum predicted number of weekly average hospitalizations increased to 192.1. The ratios between the areas under the prediction curves with and without vaccination were 0.318, indicating a 214% (CI = (192%, 250%)) increase in the number of hospitalizations when no vaccine would be present. In a comparable way, we obtained the hospitalization estimate without the Delta variant. The ratio of the areas under the two graphs (with and without the Delta variant) is 3.037, indicating a 67% (CI = (53.5%, 89.4%)) decrease in the hospitalization rate. We also tested the forecasting (forward prediction) performance of the regression model (Figure 5; Panel B). Using the data from the first two months under study, we fitted the same regression model. Then we forecasted the hospitalization rates for July and August 2021. The fitted intercept and slope coefficients (Figure 5; panel B) were 2.003 × 10^−4^ (std = 1.241 × 10^−5^) and 1.263 (std = 0.0385). As the model only used the first half data, the R-square is smaller than the full model (0.550). The model we fitted is statistically significant, and its forecasting demonstrated commendable performance. Furthermore, we conducted a regression analysis linking the hospitalization rate to wastewater concentration. The resulting slope coefficient was 0.1762 (sd= 0.0119), and an R-square value was 0.9241. Notably, the predictions from this simple regression model outperformed those of the hierarchical regression model discussed earlier. This suggests that wastewater concentration can serve as a robust predictor for forecasting hospitalization rates. Comprehensive estimation results are provided in Table S11.

**Figure 5.**
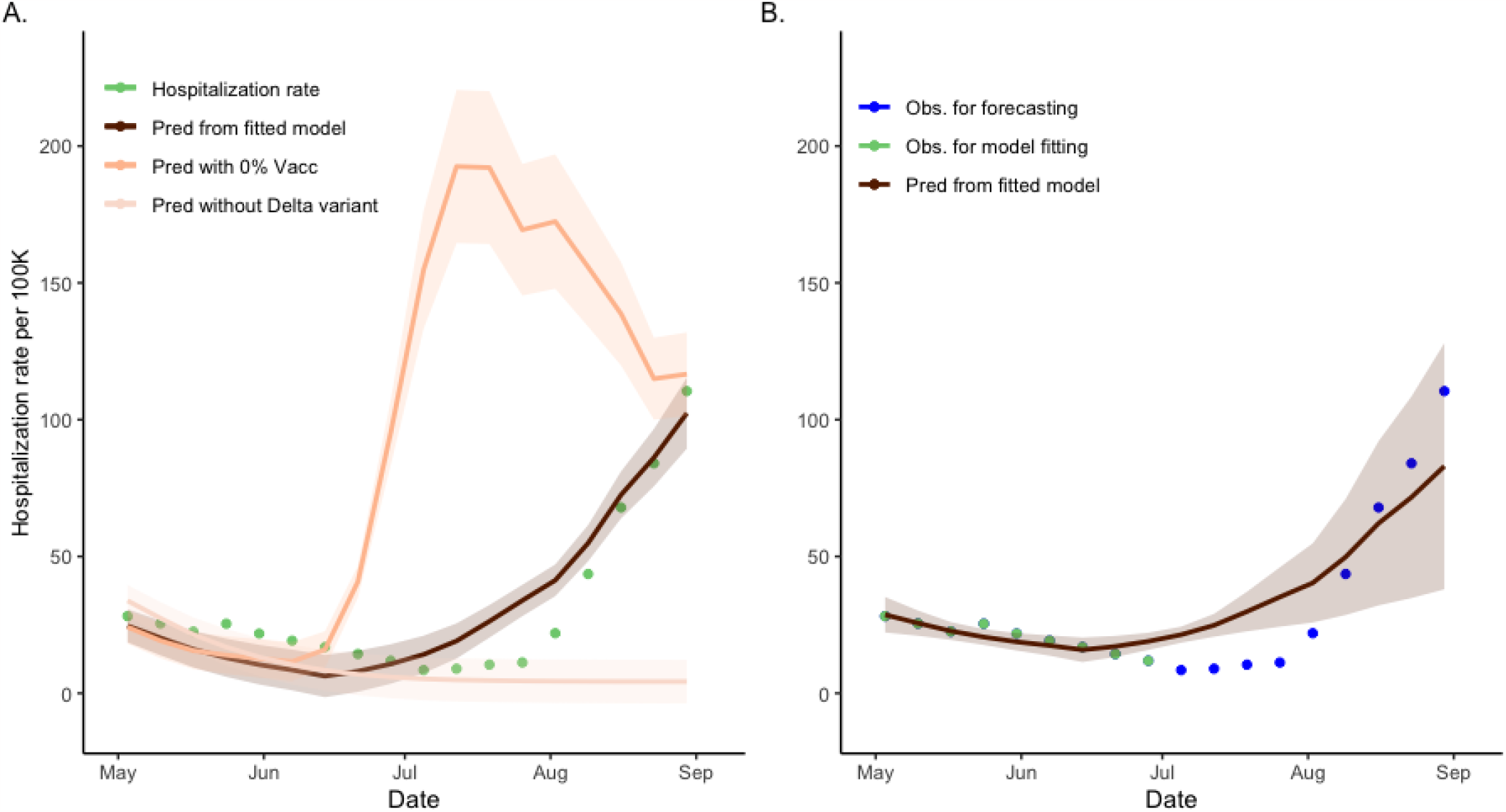
Time lag-dependent analysis of the relationship between hospitalization rate and wastewater concentration, Jefferson County, KY (USA). Panel A. Predictions and 95% confidence intervals of hospitalization rate regressed on week-lagged variables of the weekly average of wastewater concentration according to the changes of the vaccination proportion of the community. The dark line represents the prediction using the observed wastewater concentrations with 64% of community vaccination. The lighter line represents the prediction using the wastewater concentrations obtained from the model under zero community vaccination. The lightest line represents the prediction under the counterfactual modified *SVI*_2_ *RT* model with the Delta-infected model compartment zeroed-out (no Delta variant present). The green dots represent the weekly average of the observed hospitalization rate. The ratios of the areas between the prediction from the fitted model and of no vaccination are 0.318, and in the absence of the Delta variant is 3.037, respectively. **Panel B**. Forecasting (forward predictions) of mean and 95% confidence interval of hospitalization rate for July and August 2021 using a regression model fitted to data before 1 July 2021. Light blue colored dots represent the observed hospitalization rate using regression model fitting. The dark line represents the July and August 2021 forecasting using the observed wastewater concentrations with 64% of community vaccination. The dots represent the observed hospitalization rate for July and August 2021.

## 3. Discussion

The results of our large study (N = 3303) show the importance of post-vaccine environmental surveillance for the prevalence of the virus in an urban area. A major advantage of this approach is that it decreases bias implicit in publicly-available clinical case data by assessing community prevalence using antibody positivity with four waves of sequential stratified randomized sampling data. Although our work was localized to Jefferson County, where contemporaneous randomized sampling and wastewater concentration were available, it should be emphasized the model described here may be readily applicable to other locations worldwide. In addition to SARS-CoV-2, the model may be valid for other infectious diseases as well. Moreover, even though our model was run with both the SARS-CoV-2 (N1) and adjusted analysis, we noticed that the model provided reduced uncertainty with the PMMoV adjustment. Indeed, estimation of the effect of vaccination brings the related issue of refined localized model application such as high levels of tourism that may affect community vaccination levels and related observed wastewater concentrations.^8^ Here we have presented real world evidence that, in fact, small area wastewater surveillance could be used to estimate both - the effects of disease evolution as well as a community intervention, like a vaccination campaign.

It is widely recognized that even though vaccine distribution was more proportional to wealth than need, COVID-19 vaccination rates were highly effective.^18–21^ Therefore, where accessible, the impact of vaccination on community-wide prevalence of infection was readily apparent. However, for other vaccine-preventable disease, there is an urgent need for increased reliance on wastewater as a proxy for community disease impact being built from actual community level data over time, as the estimates by different methods can vary. For instance, when 90% of the student population of a college campus was vaccinated, SARS-CoV-2 in wastewater decreased^4^; but that university campus population generalization was not replicated in our community-wide survey over a longer period. In contrast to our findings, Nourbakhsh *et al*.^22^ found dissimilar trajectories from community clinical and wastewater ratios once vaccination was introduced. We suspect this difference is explained by the bias of relying on clinical data and home testing kits which became more widely available during our studied period than earlier in the pandemic and with no requirement, or in some cases option, for reporting. Whereas the Nourbakhsh *et al*.^22^ study included only publicly reported case data, the randomized selection of community participants in our study population^23^ was a comparatively less biased data source for this post-vaccine period. Our recent work has shown that even though we cannot rule out bias due to self-selection for testing, the randomized sampling approach provides better estimates of disease prevalence than administratively reported data.^23^

Our model is comparable to that used by Jiang *et al*.^24^ in that our analysis also provided estimates of prevalence; however, our estimates are based on a statistical random sample (not a clinical sample) and our regression model has a simple and explicit formula relating prevalence to observed wastewater levels of the virus. Our model further confirms the findings of Hegazy and coworkers^6^ implying the Delta variant emergence strengthened the relationship between wastewater and disease burden. Hence, our analysis provides a further proof-of-concept that our wastewater regression model could be used (after proper calibration) with other similar data to provide surrogate measures of SARS-CoV-2 prevalence in the community without the necessity for individual testing. The regression prediction correlates well with the estimated prevalence with a correlation coefficient of 0.858 (CI = (0.502, 0.975). The hospital burden findings of Wang *et al*.^25^ also compares well to our work; our results showed access to a voluntary community vaccine that reached a coverage level of 64% of the adult population decreased community hospitalizations by approximately 214%.

Yaniv *et al*.^5^ described the introduction of a new variant signal in wastewater and noted how vaccination rates and a second booster helped to control the Alpha variant, while an increase in a third booster was found to lead to a decline in Delta. When vaccination levels increase to higher coverage, hospitalizations may decline, even though the levels in wastewater remain high.^7^ Pandemic preparedness and associated public health response would benefit from new methods less dependent on continuous individual clinical testing.

Our study used five sub-county locations based on the existing wastewater infrastructure allowing observation of a small geographical area but also the aggregation of data for a countywide picture. We found that the antibody positivity varied by the sewershed. The antibody-positive individuals were lowest in sewershed MSD1 and highest in sewershed MSD3– 5 (9% for aggregate, 8% for MSD1, 9% for MSD2, and 10% for MSD3–5), indicating that previous infection may have been higher in the less dense portions of the county as compared with the urban core. Nonetheless, there are many other factors differentiating these sewershed areas that could have produced these differences. These include population sizes and demographics, or presence of stormwater or industrial discharge being combined with household sewer water. Regardless, the differences between MSD1 to 5 provide evidence of the benefit of observing results at both an aggregated and a smaller sub-county level.

For replication of our current hybrid SARS-CoV-2 model, wastewater sampling, stratified random sampling of seroprevalence, and linked vaccination data are required; the model is flexible enough to allow additional variant-specific variables. The promise of this model is that with known wastewater levels of the virus, we can predict the effect of vaccination to enable fine-tuned, and milestone-driven, public health response. The results obtained from our model, show unequivocally that the COVID-19 pandemic would have been larger and spread earlier without vaccine access. These findings provide further positive evidence for the significant role of vaccines in public health, a valuable lesson for the pandemic preparedness.

Despite its many strengths, our study has some limitations. The proportion of vaccinated respondents in the seroprevalence study was larger than the greater community (∼90% vs. 64%). Vaccine information was self-reported, and we made a simplifying assumption that the magnitude of the vaccine leakage effect is negligeable^26^ when comparing to other effects. Natural infection of a combined vaccinated and unvaccinated population (and in the absence of another way to verify vaccination) was based on antibody titers of IgG N1, an assay that has 65% sensitivity and 85% specificity^23^ a priori, with inevitable under-estimation of infection prevalence. While our serosurvey only captured adults, wastewater testing included minors. COVID-19 infected individuals can, in rare instances, shed fecal SARS-CoV-2 up to 7 months post diagnosis;^27^ viral shedding of SARS-CoV-2 can vary in relation to vaccination status and variant^28,29^ and thus was not included in our model. One of the major advances of this paper is the presentation of a relatively simple and flexible analytical model capable of using wastewater concentration to predict the effect of vaccination and new variants on prevalence and hospitalization rates. Our simulations suggest we could use as little as 50% of data to retain the statistically significant calibration conclusions (Table S12). This issue is worth further study outside the present work. Finally, the model we utilized assumed perfect protection for individuals infected with the Alpha variant against the Delta variant, as well as the insignificant seropositivity waning. While these assumptions may not be entirely valid, they appear reasonable^30^ and are unlikely to have a significant impact on our conclusions.

Overall, our work suggests that under certain conditions, it is possible to use wastewater-based epidemiology to assess both immunity acquisition in the community due to natural recovery and vaccination as well as the effect of new variants emergence and associated immune evasion to the available vaccines. The effect of vaccination on wastewater concentration as well as on community incidence of SARS-CoV-2 was substantial in Jefferson County. According to our analysis, without vaccination, one would expect about 156% of excess infections over the period of study, which corresponds to a 219% of excess wastewater concentration. The effect of the Delta variant was similarly substantial. We estimated, over the study period in Jefferson County, without Delta the amount of overall infection would decrease on average by 96% which corresponds to 88% decrease in wastewater SARS-CoV-2 (N1) normalized by PMMoV ratio. The correspondence between wastewater concentration and the number of hospitalizations was found to be strongest with the time lag for about 7 days and correlation = 0.95. Based on the regression model we estimated the effects of vaccination and variants on hospitalization rate. According to the model, without vaccination one would expect about 214% increase and without variants about 67% decrease in hospitalization rate. Using the fitted regression model for hospitalization, the predictions of hospitalization rates are at 50, 100, and 150 per 100K when SARS-CoV-2 (N1) normalized by PMMoV ratios are 0.0021, 0.0050, and 0.0077, respectively.

Our large, randomized, serosurvey suggests using the mechanistic, population level, vaccination model (*SVI*_2_ *RT*) coupled with longitudinal wastewater sampling reliably estimated the effect of vaccination on the prevalence rate in the community over the period of several months during the second and third wave of COVID-19 pandemic, in the absence of clinical data. Ours is the first study to look at a specific small area. The model can also be used to estimate the effects of vaccination and new variants emergence on the hospitalization rate and on peak hospital beds utilization, estimates critical for adequate preparedness for the next pandemic, should it arise.

## 4. Methods

### 4.1. Seroprevalence

Community-wide stratified randomized seroprevalence sampling (Table S1) was conducted in four waves from April to August 2021 in Jefferson County, Kentucky (USA) which is also the consolidated government for the city of Louisville.^23^ Seroprevalence sampling was conducted both before and during vaccination, but this analysis only considers the period after COVID-19 vaccines were made widely available to the public (N = 3,303). In some cases, due to the timing of sampling waves, respondents may have had only the first of a two-dose vaccine series. Serological positivity for nucleocapsid immunoglobulin G was used to identify participants with previous SARS-CoV-2 natural infection; vaccines used in the studied areas rely on SARS-CoV-2 viral spike protein and thus spike protein presence could be attributable to either natural infection or vaccination. Owing to elevated levels of vaccinated respondents in our study (∼90%), we only included seroprevalence measured by response to IgG N1 antibodies.^23,31^ The nucleocapsid (N1) IgG test sensitivity was 65% and the specificity was 85%.^23^ It was assumed over the study period vaccination induced antibodies do not decay below detection.

### 4.2. Levels of SARS-CoV-2 and PMMoV in the wastewater

Wastewater samples were collected twice per week from five wastewater treatment plants (N = 168; Figure S1 and Table S2) from April to August 2021. From an influent 24-hour composite sampler, 125 ml of subsample was collected and analyzed for SARS-CoV-2 (N1) and PMMoV. In a few cases due to an equipment malfunction, a grab sample was collected. The geographic area and population serviced by a wastewater treatment plant comprises a sewershed, the zone for which we consider in our model analysis across a range of population sizes, income levels and racial and ethnic diversity. Analysis used polyethylene glycol (PEG) precipitation with quantification in triplicate by reverse transcription polymerase chain reaction (RT-qPCR).^2^ Data for SARS-CoV-2 (N1) and PMMoV are reported as weekly average copies/ml of wastewater with a threshold value for SARS-CoV-2 (N1) assays of 7.5 copies/ml and for PMMoV 143 copies/ml.

### 4.3. Administrative COVID-19 data

Administrative data on COVID-19 vaccination and infected individuals’ hospitalization was provided by the Jefferson County health authority, Louisville Metro Department of Public Health and Wellness (LMPHW), under a Data Transfer Agreement. Vaccination data were geocoded to the urban sewersheds using ArcGIS Pro version 2.8.0 (Redlands, CA). Daily hospitalization data was only available aggregated at a county level.

### 4.4. Analytical model

The hybrid model for estimating the effect of vaccination and variants on longitudinal wastewater concentration was developed by combining a compartmental ecological model with a statistical linear model (Supplemental Material Appendix C). The former was used to longitudinally estimate population prevalence from the observed cross-sectional rates of seropositivity. We assumed the overall vaccination pattern as reported by the county, with the overall adult vaccination rate reaching 64%^17^ by the end of the study period. The hybrid model was used to relate the ecological model prevalence to the wastewater concentration. The ecological model, *SVI*_2_ *RT*, tracked longitudinally the proportions of individuals who were susceptible (*S*), vaccinated (*V*), infected with non-Delta variant (*I*_1_), infected with Delta variant (*I*_2_), recovered (*R*), or seropositive (*T*). We note that a version of this model that did not account for vaccination or variant was considered in our earlier work.^2^

#### 4.4.1. Regression model for wastewater concentration of SARS-CoV-2 and PMMoV ratio

Upon estimating the parameters in the _2_ model, we compared the model-calculated prevalence estimates for SARS-CoV-2 infections and vaccination levels with the wastewater concentration levels of SARS-CoV-2 (N1) and for that normalized by PMMoV.^32^ We also separately calculated two prevalence estimates according to the Alpha and Delta variants. Bayesian linear regression was performed both on the county aggregated data and stratified by sub-county wastewater treatment plant zones (sewersheds). We used the broken stick regression model to separately compare the Alpha and Delta variation effects on the wastewater concentration with regression coefficients directly. To improve the regression model stability, we used weekly average prevalence rates from the *SVI*_2_ *RT* model as the explanatory variable, and weekly aggregated average wastewater concentrations as the single outcome variable. This temporal aggregation also allowed us to use a simple posterior-profile likelihood to estimate the average change point in the broken stick regression model (see, e.g., Schwartz *et al*.^33^ for a similar approach for initial conditions imputation). We assigned non-informative priors to all regression parameters. Specifically, the non-informative Cauchy distribution was assigned to regression coefficients, and the non-informative gamma prior was assigned to the dispersion parameter error term. The regression model with intercept is used where the intercept may be interpreted as background and calibration noise related to wastewater sampling. We could see temporal differences between the Alpha and Delta variant dominant dates (Table S13), but this variability in time also considers that samples are weekly aggregated average wastewater concentrations. We did not include these variabilities of intervals in the model as the magnitudes of the observed wastewater concentration and estimated prevalence in this interval are relatively small, and model changes do not significantly alter the overall model fit.

#### 4.4.2. Estimating vaccination, variant and hospitalization effects

The strong statistical significance of the regression model relating prevalence and wastewater concentration allowed for indirect estimation of the effect of population vaccination and variants. Under the assumption the relationship between the wastewater concentration and the prevalence is not confounded by the vaccination and variants, we used the original regression equation derived from the collected wastewater and seroprevalence data to estimate the wastewater concentration over time. To estimate the vaccination effect, we compared these concentrations with hypothetical ones obtained when the vaccination term was zeroed out in the *SVI*_2_ *RT*model. In a comparable manner, we estimated the effect of the introduction of the Delta variant. Finally, we performed the longitudinal, regression-based analysis relating the community hospitalization to observed wastewater concentrations. In the three analyses we quantified the effects by calculating the size of the effects relative to the factual (observed) states.

#### 4.4.3. Competing risk model with two different virus variants

Wastewater samples were prepared for whole genome sequencing,^14,34^ and the proportion of observed SARS-CoV-2 variants was estimated for each sewershed based on variant dominance (Table S13). Two variants were present in the study area during the study period: Alpha was dominant from April until July, while Delta was dominant from July until August.^14,34^ To reflect the infections before and after the emergence of the Delta variant, we incorporated into our *SVI*_2_*RT* model the two different infection compartments (*I*_1_ and *I*_2_) reflecting both the infection competition and temporal heterogeneity caused by two different variants of the virus.

### 4.5. Ethics

For the seroprevalence and data provided by the LMPHW under a Data Transfer Agreement, the University of Louisville Institutional Review Board approved this as Human Subjects Research (IRB number: 20.0393). For the wastewater data, the University of Louisville Institutional Review Board classified this as non-human subjects research (reference #: 717950).

## Data availability

The seroprevalence, wastewater levels, and hospitalization information data used in the study can be accessed from the website https://github.com/cbskust/DSA_Seroprevalence. The computer code that implemented our model-based analysis will be made available immediately after publication.

## Acknowledgements

We thank the Louisville/Jefferson County Metropolitan Sewer District for their valuable collaboration in wastewater sample collection. This study was supported by Centers for Disease Control and Prevention (75D30121C10273), Louisville Metro Government, James Graham Brown Foundation, Owsley Brown II Family Foundation, Welch Family, Jewish Heritage Fund for Excellence, the National Institutes of Health, (P20GM103436), the Rockefeller Foundation, and the National Sciences Foundation (DMS-2027001).

## Author contributions

AB, TS, KEP, and RK conceived and developed the idea for the study. KEP supervised the serology assays; ABS and KK performed the serology assays, performed quality control assessments, and uploaded the data to the redcap database. TS and RH supervised the wastewater analyses. GAR and BC performed the model-based data analyses. RHH and GAR wrote the first draft of the manuscript with all authors contributing to the interpretation of data and critical revision of the article. MB accessed and verified the data reported in the study. All authors had full access to all the data in the study and had final responsibility for the decision to submit for publication.

## Supplementary information

## Appendix A. SARS-CoV-2 seroprevalence by wave and sewershed, Jefferson County, KY (USA)

**Table S1.**
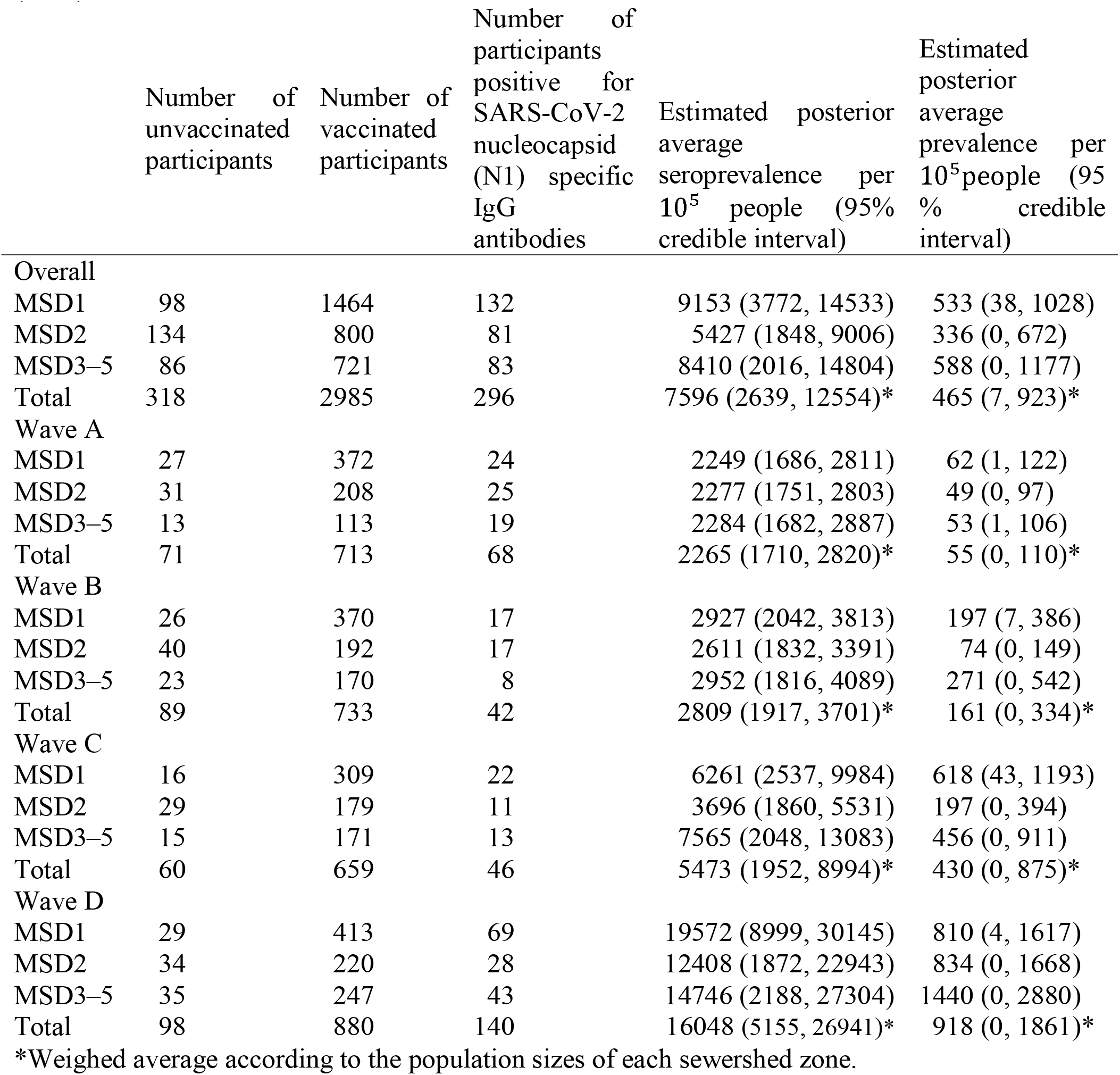
SARS-CoV-2 seroprevalence by wave and sewershed, Jefferson County, KY (USA).

|  | Number of<br>unvaccinated<br>participants | Number of<br>vaccinated<br>participants | Number of<br>participants<br>positive for<br>SARS-CoV-2<br>nucleocapsid<br>(N1) specific<br>IgG<br>antibodies | Estimated posterior<br>average<br>seroprevalence per<br>10<sup>5</sup> people (95%<br>credible interval) | Estimated<br>posterior<br>average<br>prevalence per<br>10<sup>5</sup> people (95%<br>credible<br>interval) |
| --- | --- | --- | --- | --- | --- |
| Overall |  |  |  |  |  |
| MSD1 | 98 | 1464 | 132 | 9153 (3772, 14533) | 533 (38, 1028) |
| MSD2 | 134 | 800 | 81 | 5427 (1848, 9006) | 336 (0, 672) |
| MSD3–5 | 86 | 721 | 83 | 8410 (2016, 14804) | 588 (0, 1177) |
| Total | 318 | 2985 | 296 | 7596 (2639, 12554)* | 465 (7, 923)* |
| Wave A |  |  |  |  |  |
| MSD1 | 27 | 372 | 24 | 2249 (1686, 2811) | 62 (1, 122) |
| MSD2 | 31 | 208 | 25 | 2277 (1751, 2803) | 49 (0, 97) |
| MSD3–5 | 13 | 113 | 19 | 2284 (1682, 2887) | 53 (1, 106) |
| Total | 71 | 713 | 68 | 2265 (1710, 2820)* | 55 (0, 110)* |
| Wave B |  |  |  |  |  |
| MSD1 | 26 | 370 | 17 | 2927 (2042, 3813) | 197 (7, 386) |
| MSD2 | 40 | 192 | 17 | 2611 (1832, 3391) | 74 (0, 149) |
| MSD3–5 | 23 | 170 | 8 | 2952 (1816, 4089) | 271 (0, 542) |
| Total | 89 | 733 | 42 | 2809 (1917, 3701)* | 161 (0, 334)* |
| Wave C |  |  |  |  |  |
| MSD1 | 16 | 309 | 22 | 6261 (2537, 9984) | 618 (43, 1193) |
| MSD2 | 29 | 179 | 11 | 3696 (1860, 5531) | 197 (0, 394) |
| MSD3–5 | 15 | 171 | 13 | 7565 (2048, 13083) | 456 (0, 911) |
| Total | 60 | 659 | 46 | 5473 (1952, 8994)* | 430 (0, 875)* |
| Wave D |  |  |  |  |  |
| MSD1 | 29 | 413 | 69 | 19572 (8999, 30145) | 810 (4, 1617) |
| MSD2 | 34 | 220 | 28 | 12408 (1872, 22943) | 834 (0, 1668) |
| MSD3–5 | 35 | 247 | 43 | 14746 (2188, 27304) | 1440 (0, 2880) |
| Total | 98 | 880 | 140 | 16048 (5155, 26941)* | 918 (0, 1861)* |

\*Weighed average according to the population sizes of each sewershed zone.

## Appendix B. Studied wastewater treatment plant zones (sewersheds), Jefferson County, KY (USA)

**Figure S1.**
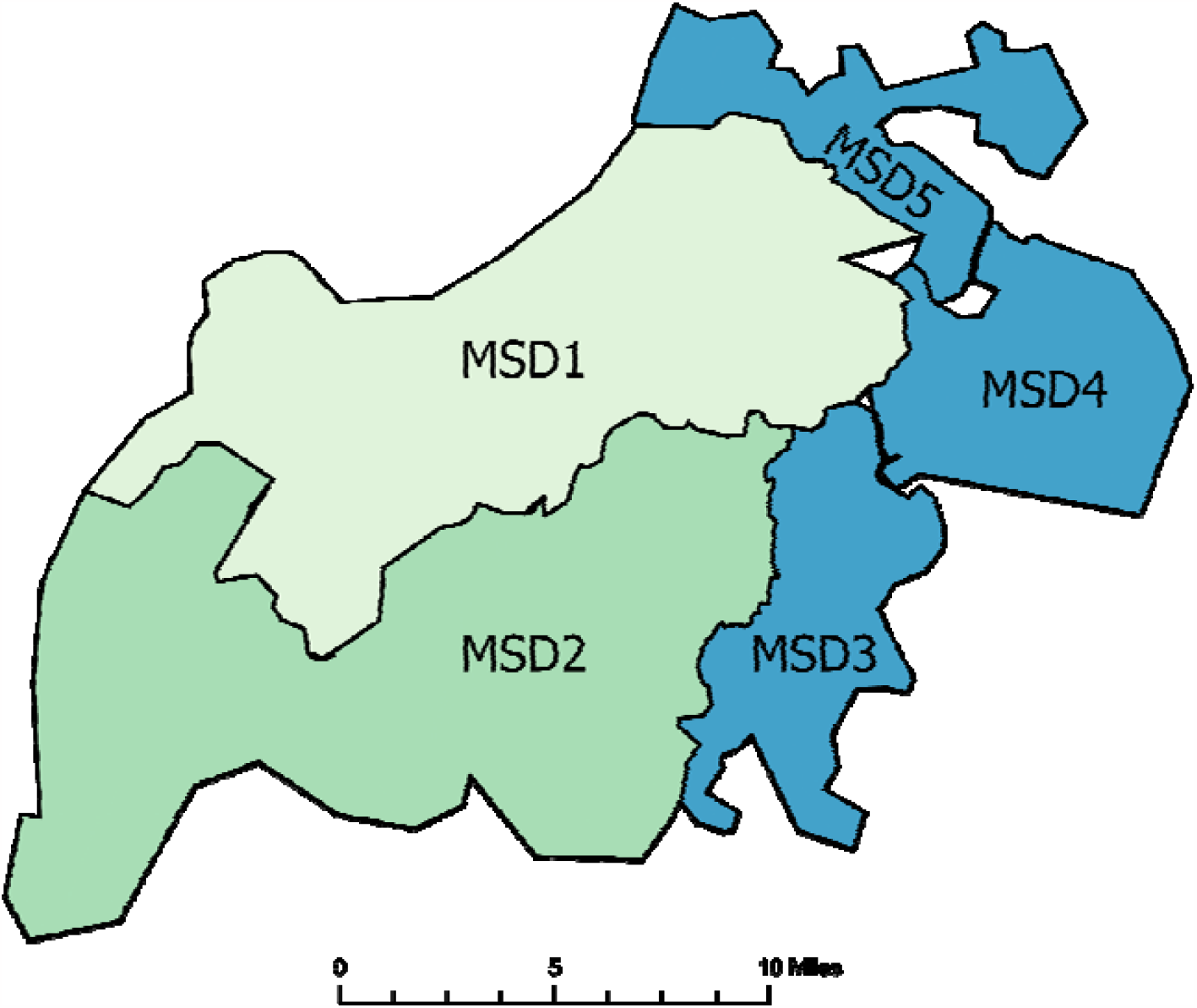
Studied wastewater treatment plant sewersheds, Jefferson County, Kentucky (USA).

**Table S2.** Characteristics of studied wastewater treatment plant sewersheds of Jefferson County, KY (USA).

| **Sewershed** | **Income**<br>**(USD\$)<sup>a</sup>** | **Population<sup>a</sup>** | **Area**<br>**(km<sup>2</sup>)** | **Combined**<br>**sewer<sup>b</sup>** |
| --- | --- | --- | --- | --- |
| MSD1<br>Morris Forman Water Quality Treatment Center | 54,138 | 349,850 | 280 | Yes |
| MSD2<br>Derek R. Guthrie Water Quality Treatment Center | 53,577 | 295,910 | 332 | No |
| MSD3<br>Cedar Creek Water Quality Treatment Center | 76,606 | 55,928 | 80 | No |
| MSD4<br>Floyds Fork Water Quality Treatment Center | 113,699 | 32,460 | 88 | No |
| MSD5<br>Hite Creek Water Quality Treatment Center | 106,769 | 31,269 | 67 | No |

<sup>a</sup>Based on 2018 U.S Census Bureau American Community Survey (ACS) block group data aggregated to the wastewater catchment areas with overlapping block group centroids. Income is mean median household. For more on wastewater site selection see Yeager et al., 2021.
<sup>b</sup>Combined sewers include wastewater and stormwater and SARS-CoV-2 concentrations may be expected to fluctuate more as a result.

## Appendix C. Population vaccination model (SVI_2_RT)

The equation shown in (1) describes the time-evolution of the proportions of individuals who are susceptible (*S*), vaccinated (*V*), infected with Alpha variant (*I*_1_), infected with Delta variant (*I*_2_) removed (*R*), and seropositive (*T*). We assume the total initial population of susceptibles is large with a small initial fraction of infected. The model equations are:

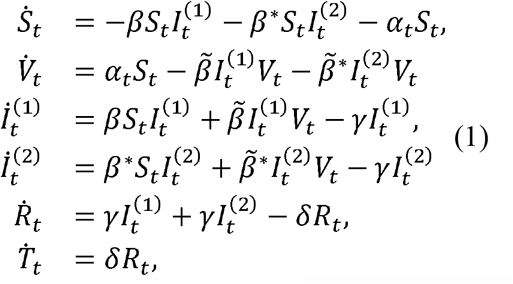

with the initial condition 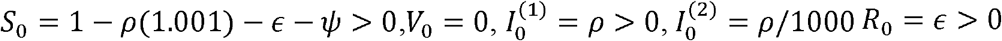, and *T*_0 = ψ< 0;_

Here,β and 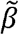 are the rates of infection of respectively, unvaccinated and vaccinated, and β* and 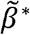 are the rates of infection according to Delta variant. As our compartment model has two infection compartments, it is called the variant competition model.^1^ The observed data in this analysis do not have any information about infection from the Delta variant, and an increase in the number of parameters makes model estimation difficult and may lead to identifiability problems. So, we set β^*^ and 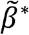 at the values 50% higher than βand 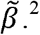 The function of α_*t*_ represents a changing rate of vaccination over time. The vaccination process may be changed according to a policy or vaccine supply, so we set the vaccination rate α_t_ to match the empirical percentage of the vaccinated population in Jefferson County at the end of August 2021. Additionally, γ is the rate of recovery, and δ is the rate at which antibodies build to a detectable level after recovery. The *SVI*_2_ *RT* model parameters to be estimated are given by the vector θ = (β,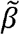, γ, δ, ρ,ϵ, ψ).

To obtain the serial estimates of incidence and prevalence from the observed seropositivity levels in four waves of testing, we adapt the idea of an ODE-based survival model proposed recently.^3,4^ According to that model, the scaled quantities 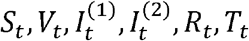 may be considered as respective probabilities of a randomly selected individual in a large population, being either susceptible, vaccinated, infected with different virus variant, recovered, or seroprevalent at time *t*. Consequently, we consider the results *Z*(*t*) of all individual antibody-based tests conducted at times *t* as independent Bernoulli variables:

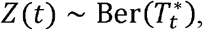

where 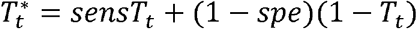 is the specificity adjusted probability of a positive test. For our analysis, both *sens* and *spe*, are additional parameters to be estimated. We assigned the informative priors to *sens*, and *spe*, from available clinical data.

Assuming at time *t, n*_t_ individuals are tested with _t_ having positive results, the corresponding log-likelihood function is:

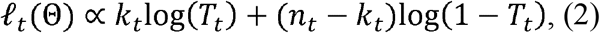

where Θ = (β,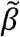, γ, δ, ρ,ϵ, ψ, *spe, sens*). is the vector of parameters to be identified. Given the testing data at m≥1 time points *t*_1_,…, *t*_*m*_, we then aim to find parameter values θ that maximizes the posterior log-likelihood function:

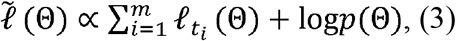

where *p*( Θ) is the prior distribution on Θ to be determined from our previous work^3,4^. Hence, we seek the values of Θ that maximize our posterior log-likelihood function (3). The entire system (1) must be solved for each parameter combination.

### S3.1. Incidence, prevalence, and seroprevalence estimation

Posterior serial estimates of the relative rates of incidence, prevalence, and seropositivity were obtained from the *SVI*_2_ *RT* model as the time-dependent vector:

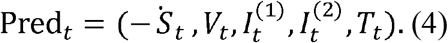

Here 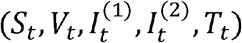 is the family of trajectories of (1) evaluated at the posterior distribution of the vector Θ. In practice, the distribution of Predt is approximated by taking a random sample of size m from the converged MCMC sampler. In our case *m* = 2000. To obtain daily incidence rates (Incd) we have used the approximation 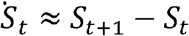 and consequently took Incd =sd − *S*_d+1_ where *d* corresponds to a specific day of interest. The estimated prediction counts were obtained by multiplying the rates in Pred by the appropriate population numbers.

**Table S3.**
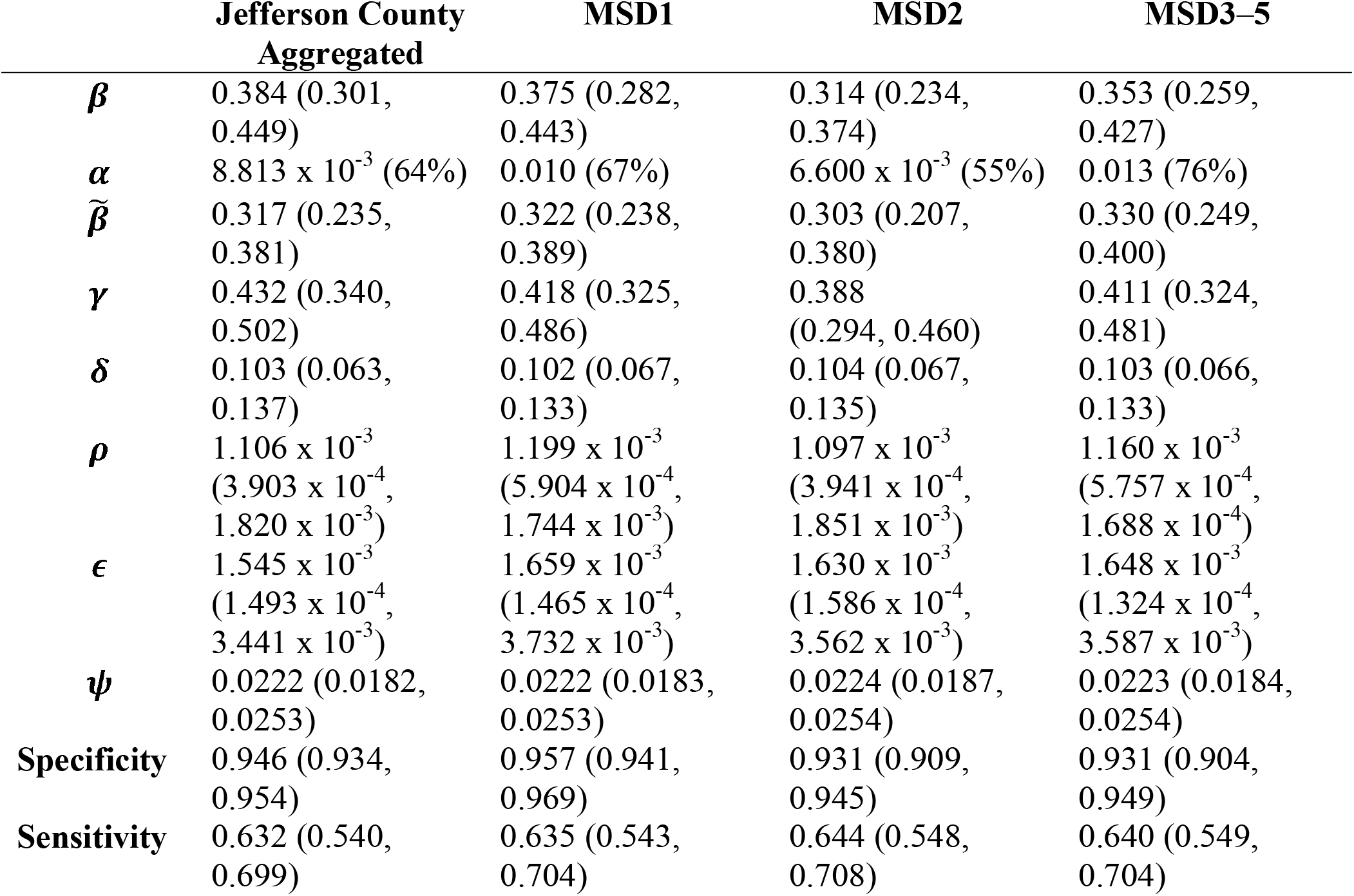
Posterior mean estimates of the *SVI*_2_ *RT* model parameters in sewersheds of Jefferson County, KY (USA). The area-specific Hamiltonian Markov chain Monte Carlo (MCMC) posterior estimates are based on seropositivity data aggregated across Jefferson County and stratified by sewersheds. The corresponding 95% credible bounds are provided in parenthesis. The results are based on MCMC implemented via *Rstan* library, with a 6000- and 2000-step burn-in.

**Table S4.**
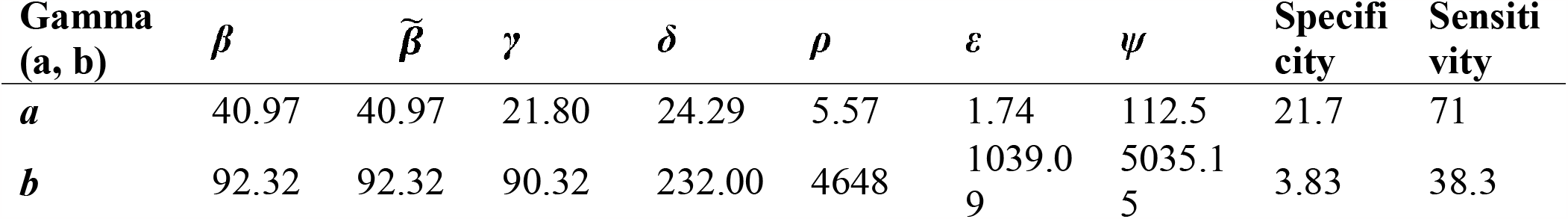
The prior distribution specifications for the *SVI*_2_ *RT* model. Parameters were given Gamma prior distributions, with hyper-parameters (*a, b*) listed in the table below.

**Table S5.** Summary of the Bayesian broken stick regression results in sewersheds of Jefferson County, KY (USA). Dispersion (σ)is the standard deviation of the error term of the linear regression.

| Sewershed | Parameters | Linear regression model |
| --- | --- | --- |
|  |  | Posterior mean (95% credible interval) |
| Jefferson County Aggregated | Intercept | $-4.222 \times 10^{-4}$ ($-9.458 \times 10^{-4}$, $7.921 \times 10^{-5}$) |
|  | Alpha variant | 0.815 (-0.023, 1.717) |
|  | Delta variant | 0.385 (0.318, 0.455) |
| | Dispersion ($\sigma$) | $6.483 \times 10^{-4}$ ($4.543 \times 10^{-4}$, $9.490 \times 10^{-4}$) |
| MSD1 | Intercept | $-7.012 \times 10^{-4}$ ($-1.385 \times 10^{-3}$, $1.493 \times 10^{-5}$) |
|  | Alpha variant | 1.126 (0.096, 2.112) |
|  | Delta variant | 0.240 (0.181, 0.296) |
| | Dispersion ($\sigma$) | $8.153 \times 10^{-4}$ ($5.739 \times 10^{-4}$, $1.186 \times 10^{-3}$) |
| MSD2 | Intercept | $-2.099 \times 10^{-4}$ ($-8.447 \times 10^{-4}$, $4170 \times 10^{-4}$) |
|  | Alpha variant | 0.881 (-0.325, 2.073) |
|  | Delta variant | 0.557 (0.482, 0.631) |
| | Dispersion ($\sigma$) | $8.939 \times 10^{-4}$ ($6.330 \times 10^{-4}$, $1.300 \times 10^{-3}$) |
| MSD3–5 | Intercept | $-2.963 \times 10^{-4}$ ($-7.426 \times 10^{-4}$, $1.508 \times 10^{-4}$) |
|  | Alpha variant | 0.630 (-0.155, 1.434) |
|  | Delta variant | 0.201 (0.163, 0.240) |
| | Dispersion ($\sigma$) | $5.635 \times 10^{-4}$ ($3.961 \times 10^{-4}$, $8.323 \times 10^{-4}$) |

**Table S6.** Sensitivity analysis. The transmission rates of the Delta variant, denoted as β*, set to 120%, 150%, 200%, 250%, and 300% of the transmission rate of the Alpha variant, denoted as β. The second column represents the corresponding increases in the basic reproduction numbers.

| **Increasing amount of transmission rate**<br>**of Delta variant** |  |
| --- | --- |
|  | **$R_0$** |
| 1.2 | 1.06 |
| 1.5 | 1.33 |
| 2.0 | 1.78 |
| 2.5 | 2.22 |
| 3.0 | 2.67 |

**Table S7.**
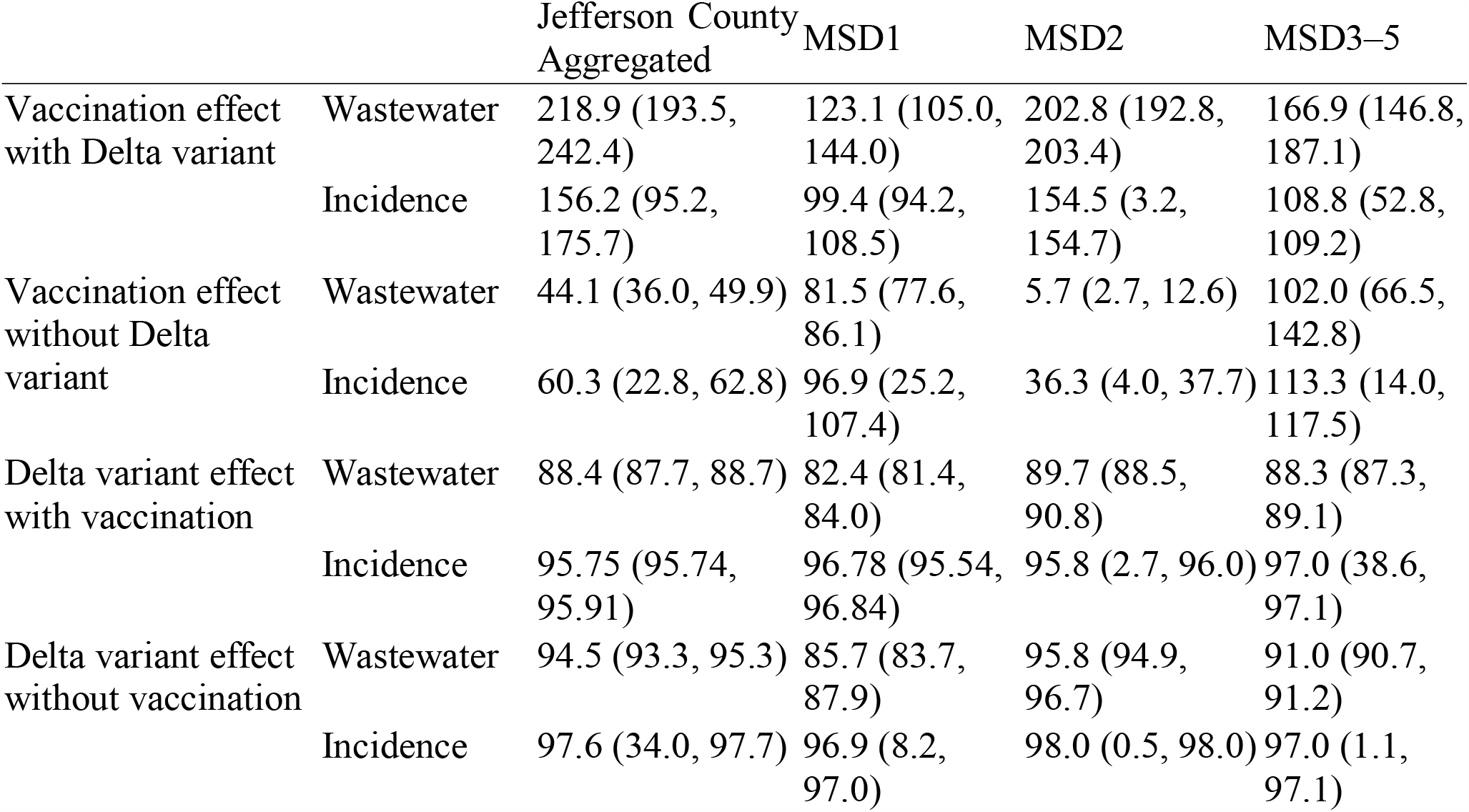
Summary of the effects of the vaccination and Delta variant in sewersheds of Jefferson County, KY (USA). Percentage reduction due to vaccination effect or excess due to Delta variant on estimates of wastewater concentration and incidence rate. In parenthesis, we give lower and upper bounds of 95% credible interval.

**Table S8.** Summary of the effects of the vaccination and Delta variant in sewersheds of Jefferson County, KY (USA). The absolute values of difference between estimated wastewater concentrations and incidences due to vaccination effect or Delta variant. In parenthesis, we give lower and upper bounds of the 95% credible interval.

|  |  | Jefferson<br>County<br>Aggregated | MSD1 | MSD2 | MSD3–5 |
| --- | --- | --- | --- | --- | --- |
| Vaccination effect<br>with Delta variant | Wastewater | 0.419 (0.377,<br>0.463) | 0.266 (0.236,<br>0.296) | 0.486 (0.456,<br>0.518) | 0.250 (0.223,<br>0.277) |
|  | Incidence | 0.410 (0.035,<br>0.837) | 0.382 (0.033,<br>0.797) | 0.301 (0.000,<br>0.603) | 0.413 (0.001,<br>0.826) |
| Vaccination effect<br>without Delta<br>variant | Wastewater | 0.010 (0.008,<br>0.013) | 0.033 (0.030,<br>0.035) | 0.002 (0.001,<br>0.004) | 0.019 (0.014,<br>0.024) |
|  | Incidence | 0.006 (0.000,<br>0.012) | 0.011 (0.000,<br>0.023) | 0.003 (0.000,<br>0.005) | 0.012 (0.000,<br>0.025) |
| Delta variant effect<br>with vaccination | Wastewater | 0.182 (0.180,<br>0.185) | 0.190 (0.183,<br>0.197) | 0.233 (0.230,<br>0.237) | 0.142 (0.140,<br>0.143) |
|  | Incidence | 0.246 (0.035,<br>0.456) | 0.372 (0.033,<br>0.711) | 0.187 (0.000,<br>0.374) | 0.362 (0.001,<br>0.724) |
| Delta variant effect<br>without vaccination | Wastewater | 0.583 (0.541,<br>0.628) | 0.440 (0.412,<br>0.470) | 0.654 (0.625,<br>0.684) | 0.384 (0.362,<br>0.406) |
|  | Incidence | 0.642 (0.001,<br>1.82) | 0.739 (0.000,<br>1.478) | 0.486 (0.000,<br>0.972) | 0.767 (0.000,<br>1.534) |

**Table S9.** Vaccination effect of the incidence estimation of Jefferson County, KY (USA). The absolute values of differences between cumulative number of the estimated incidences due to vaccination effect or Delta variant effect0. In parenthesis, we give lower and upper bounds of 95% credible interval. For comparison between sewershed zone, we estimated the incidence per 10^5^ population.

|  | Jefferson County<br>Aggregated | MSD1 | MSD2 | MSD3–5 |
| --- | --- | --- | --- | --- |
| Incidence Vaccination | 40,085 (3,507,<br>83,678) | 38,205 (3,264,<br>79,673) | 30,146 (1,<br>60,293) | 40,663 (70,<br>81,361) |
| Delta variant | 24,567 (3,534,<br>45,601) | 37,210 (3,309,<br>71,111) | 18,693 (1,<br>37385) | 36,210 (70,<br>72,351) |

**Table S10.** Correlation coefficients and 95% credible intervals between wastewater concentration and the estimated prevalence from the Alpha variant mutation in sewersheds of Jefferson County, KY (USA).

|  | Jefferson County<br>Aggregated | MSD1 | MSD2 | MSD3–5 |
| --- | --- | --- | --- | --- |
| Incidence Vaccination | 0.51185 (-<br>0.3296, 0.9427) | 0.5773 (-0.2731, 0.9431) | 0.9105 (0.6217, 0.9914) | 0.1243 (-0.6859, 0.8364) |

**Table S11.** Simple linear regression model for the hospitalization rate on the observed weekly average of wastewater concentration.

| Response | Parameters | Estimate | Std. | t statistic | P-value |
| --- | --- | --- | --- | --- | --- |
| Hospitalization<br>rate | Intercept | 1.284<br>$\times 10^{-4}$ | 2.729<br>$\times 10^{-5}$ | 4.705 | 0.0002 |
|  | Wastewater<br>concentration | 0.1762 | 0.0119 | 14.835 | 0.0000 |

**Table S12.** A simulation study summary for hierarchical regression. Each regression model was fitted using random sample data. Sample portions considered are: 100%, 83% 67%, 50%, and 33%.

| Percentage | $R^2$ | F statistics | P-value |
| --- | --- | --- | --- |
| 100 | 0.9000 | 145.5 | $9.273 \times 10^{-10}$ |
| 83 | 0.8842 | 99.26 | $1.878 \times 10^{-7}$ |
| 67 | 0.8411 | 52.93 | $2.679 \times 10^{-5}$ |
| 50 | 0.7735 | 23.90 | $1.775 \times 10^{-3}$ |
| 33 | 0.2095 | 1.06 | 0.3614 |

**Figure S2.**
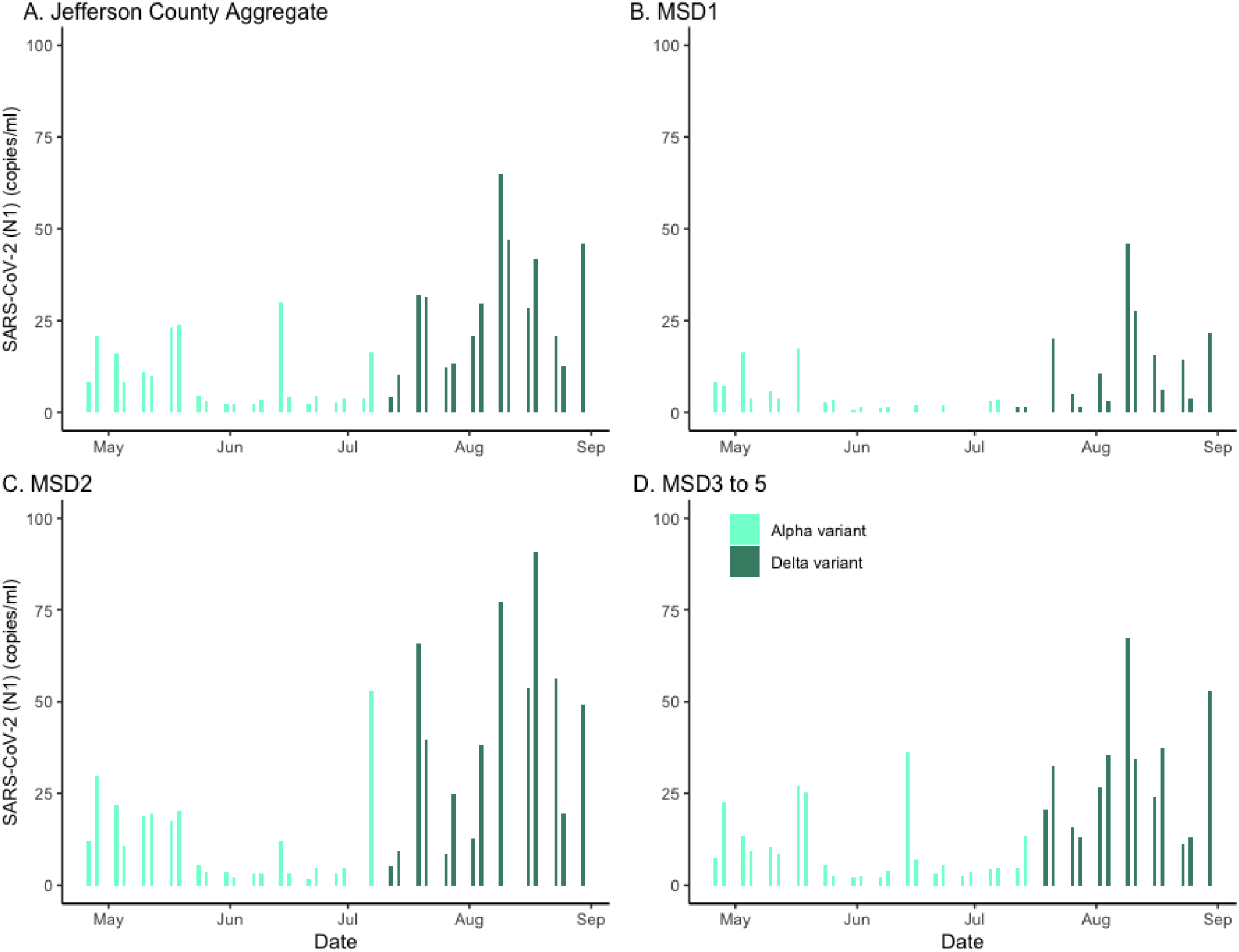
SARS-CoV-2 (N1) wastewater concentration in sewersheds of Jefferson County, KY (USA). The wastewater concentrations during Alpha and Delta variants are represented in bars (light green for Alpha variant, dark green for Delta variant). The panels compare aggregated concentration for Jefferson County (Panel A) as well as stratified by sewershed (Panels B–D).

**Figure S3.**
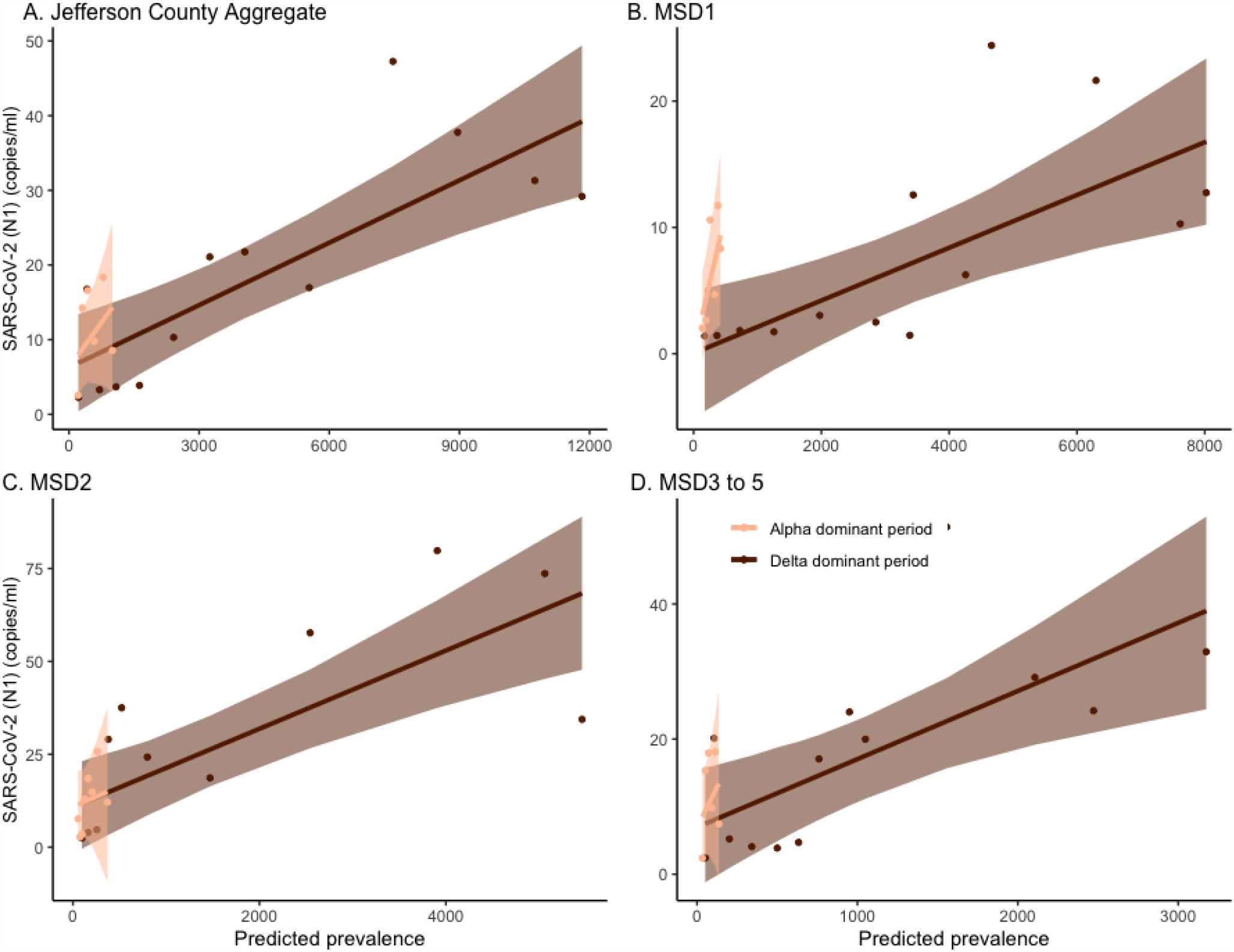
Prevalence versus SARS-CoV-2 (N1) wastewater concentration in sewersheds of Jefferson County, KY (USA). Bayesian regression between predicted weekly prevalence of SARS-CoV-2 infections from the Alpha and Delta variants and wastewater in the entire Jefferson County (Panel A) as well as stratified by sewershed (Panels B–D). The darker straight line is the fitted Bayesian regression line for the Delta variant. The darker shade marks the 95% credible interval: the lighter line and shade mark for the Alpha variant. The data points for the Alpha variant are minor (6 for Panels A, B, and D, 8 for C).

**Figure S4.**
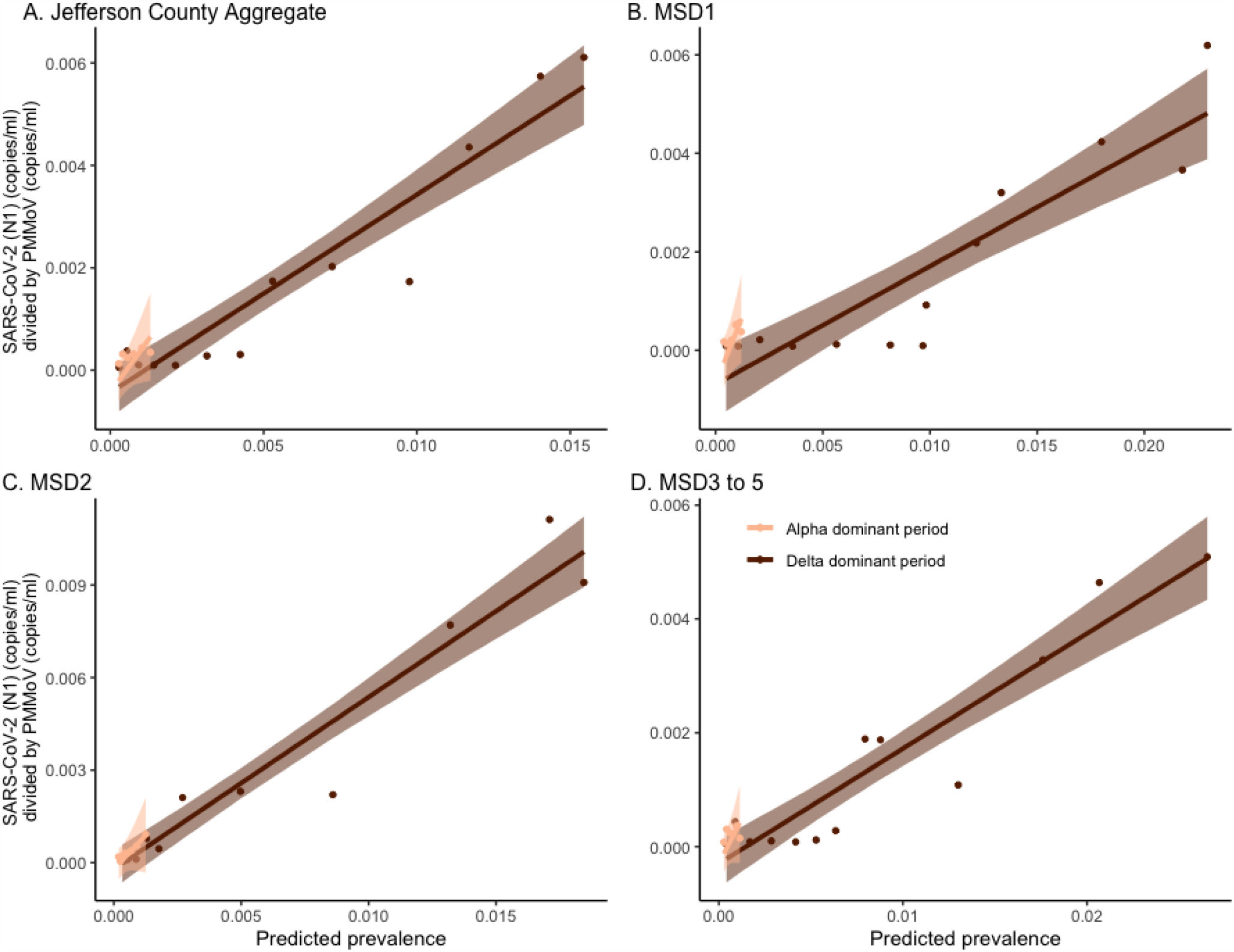
Prevalence versus wastewater SARS-CoV-2 (N1) normalized by pepper mild mottle virus concentration in sewersheds of Jefferson County, KY (USA). Bayesian regression between predicted weekly prevalence of SARS-CoV-2 infections from the Alpha and Delta variants and wastewater in the entire Jefferson County (Panel A) as well as stratified by sewershed (Panels B–D). The darker straight line is the fitted Bayesian regression line for th Delta variant. The darker shade marks the 95% credible interval: the lighter line and shade mark for the Alpha variant. The data points for the Alpha variant are very few (6 for Panels A, B, and D, 8 for C).

**Figure S5.**
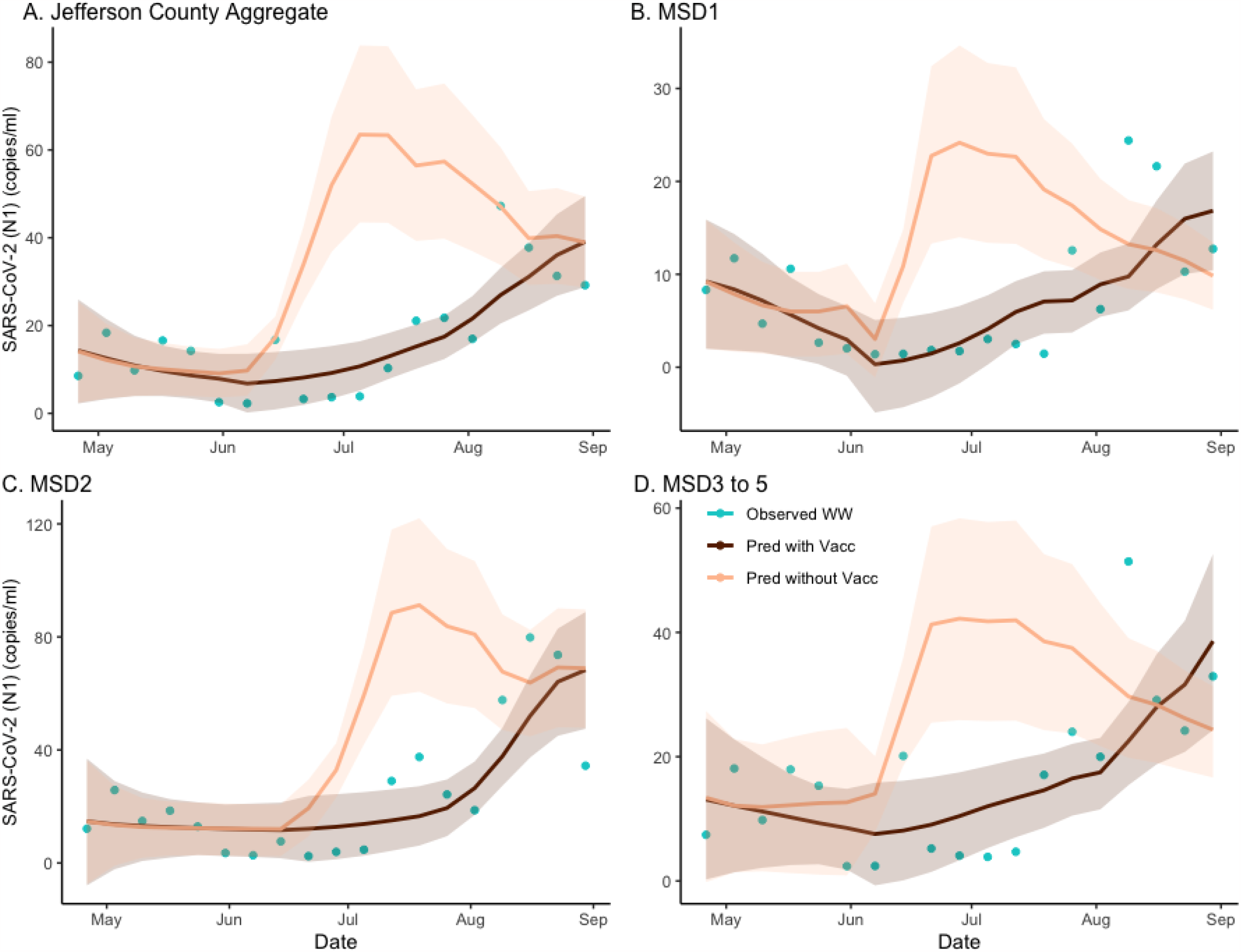
The estimated effect of vaccination on SARS-CoV-2 (N1) wastewater concentration in sewersheds of Jefferson County, KY (USA). The dark brown line is the regression-based fit to the wastewater concentration and the light brown line is the prediction of wastewater concentration using synthetic prevalence from the model with the Delta variant effect zeroed out. The shaded areas represent 95% credible intervals. The blue dots are observed weekly average wastewater concentration. The panels compare the variant effect on wastewater concentration for Jefferson County (Panel A) as well as stratified by sewershed (Panels B–D).

**Figure S6.**
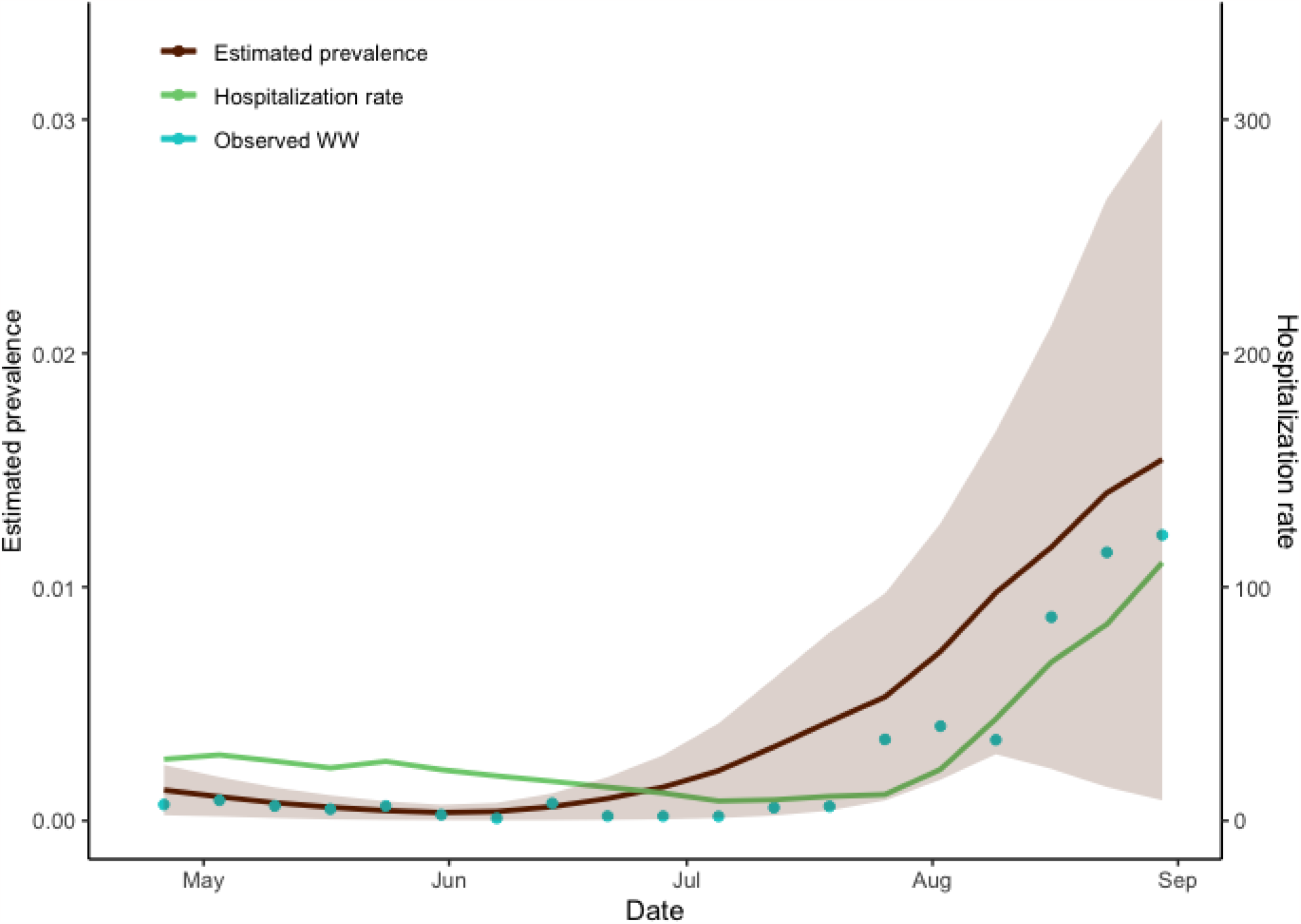
SARS-CoV-2 prevalence and hospitalizations versus SARS-CoV-2 (N1) wastewater concentration normalized by pepper mild mottle virus, Jefferson County, KY (USA). Relationship among observed wastewater concentration, the hospitalization rate, and estimated prevalence. The dark brown line represents the estimated prevalence, and the shaded area is the 95% credible interval of MCMC simulation. The green line is the weekly average of daily hospitalization rate of Jefferson County, and the blue dots represent the weekly average of wastewater concentrations. The Pearson correlation coefficient of estimated prevalence and wastewater concentration is 0.858 (95% CI = (0.502, 0.975)) and that of hospitalization rate and wastewater concentration is 0.722 (95% CI = (0.216, 0.955)).

**Figure S7.**
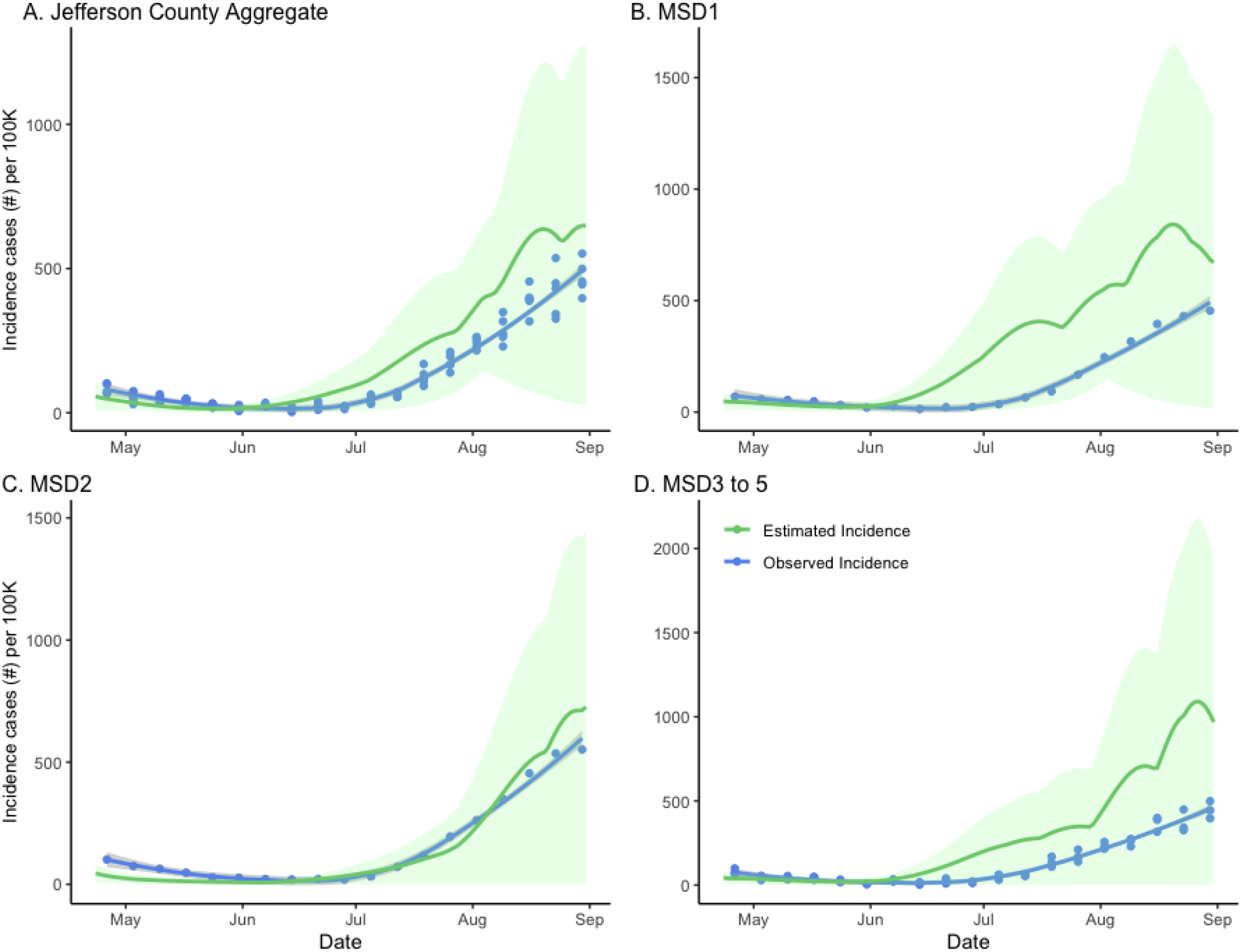
Clinical versus estimated incidence in sewersheds of Jefferson County, KY (USA). Posterior density and credibility bounds (green curve) of the weekly aggregated incidence rate as predicted by the model compared to official weekly incidence for Jefferson County (blue dots and trend line) as reported by the Jefferson County Health Department. The model plots are based on Hamiltonian MCMC samples, with 6000 steps and 2000 steps burn-in period. The panels compare aggregated incidence for Jefferson County (Panel A) as well as stratified by sewershed (Panels B–D).

**Figure S8.**
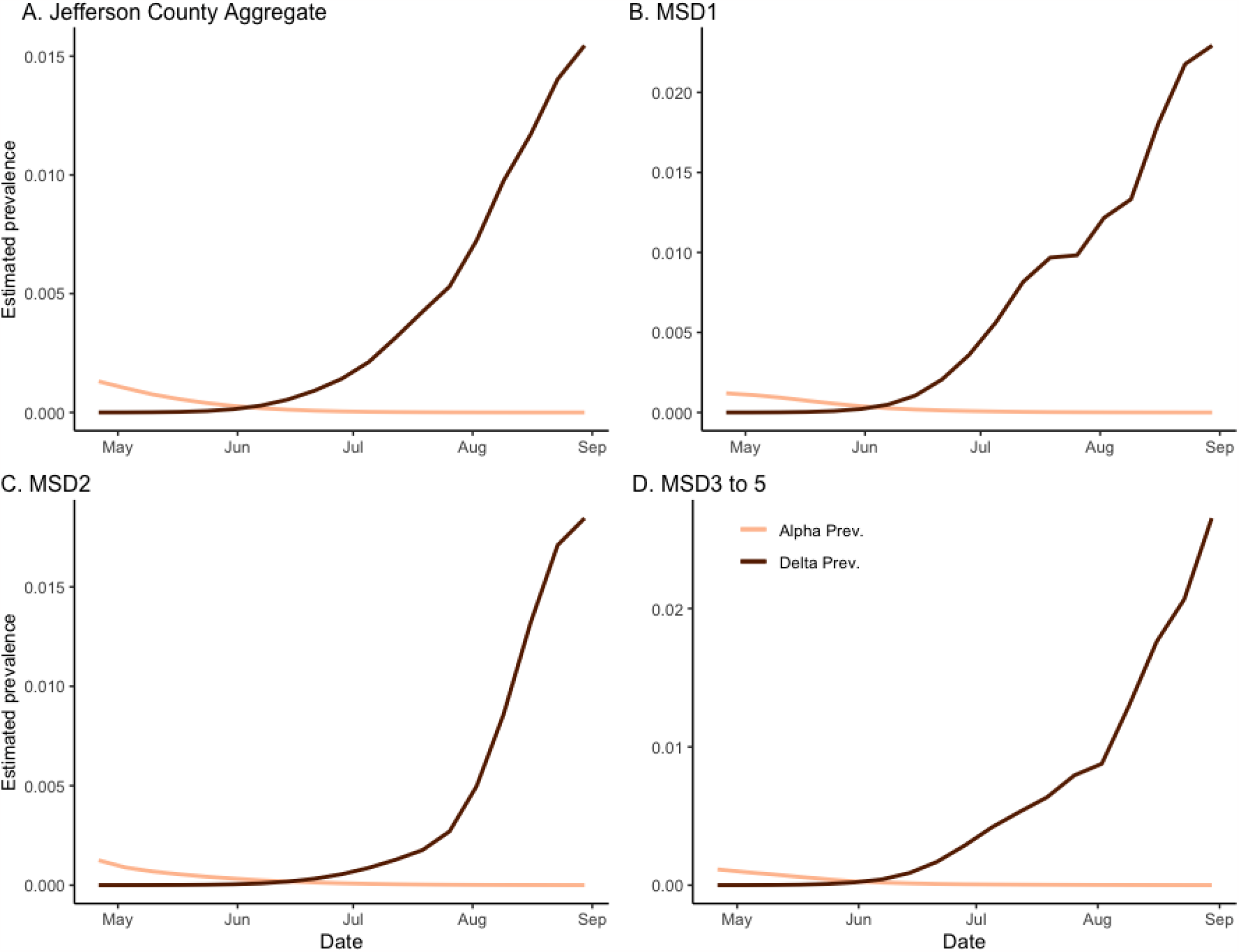
Estimated prevalence for SARS-CoV-2 Alpha and Delta variants in sewersheds of Jefferson County, KY (USA). Estimated prevalence of the Alpha and Delta variants by the model. Two estimated prevalence lines crosses at 5 June 2021 (for Panels A, B, and D) and 15 June 2021 (for Panel C), corresponding to the middle of the period of the Alpha variant being dominant. The panels compare prevalence for Jefferson County (Panel A), as well a stratified by sewershed (Panels B–D).

**Figure S9.**
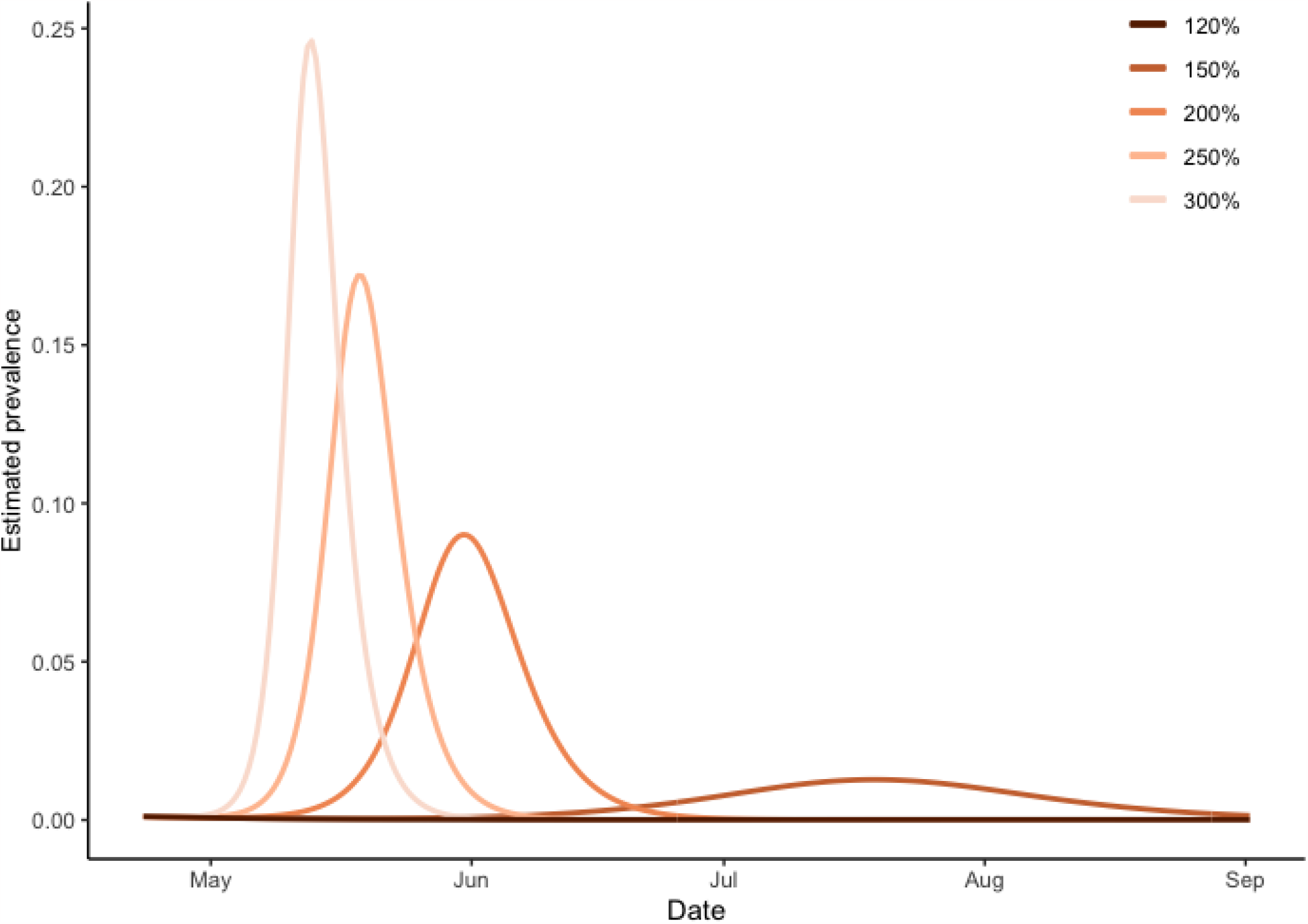
Sensitivity analysis for change of prevalence according to the change of transmission rate for Delta variant. The amount of the transmission rate for Delta variant are set from 120% to 300% which is as large as the Alpha variant transmission rate. The corresponding basic reproduction numbers are seen to change from 1.3 for 120% to 3.2 for 300%.

### S3.2. Details on regression model for wastewater concentration

To relate the *SVI*_2_ *RT* model predictions to the serial wastewater measurements of SARS-CoV-2 (N1) concentrations and normalized SARS-CoV-2 (N1) divided by pepper mild mottle virus (PMMoV) ratio, the Bayesian linear regressions were performed based on aggregated county data and data stratified by sewershed area.

To obtain the broken stick linear regression models^5^, the procedure was as follows: Let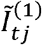 and 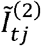 be the model estimated percentage prevalence corresponding to the same week and sewershed area for the Alpha and Delta variants, respectively. We first define two basic functions *B*_*l*_(*tj*) and *B*_*r*_(*tj*):

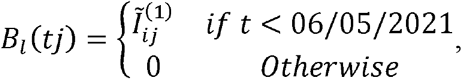

and

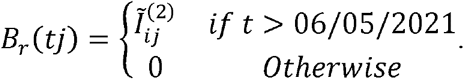

*W*_*tj*_ represents the weekly aggregated average wastewater concentration. We can now fit the model of the form:

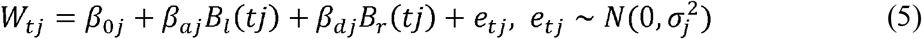

In the Bayesian linear regression models, non-informative priors were assigned. Specifically, the non-informative Cauchy distribution was assigned to the regression coefficients, and the non-informative gamma prior was assigned to the dispersion parameter of the error term.

### S3.3. Time lag-dependency between wastewater concentration and hospitalization rate

It takes a certain period for the patient to be admitted to the hospital to receive treatment. To identify the time lag-dependency between wastewater concentration and hospitalization rate, a simple linear regression analysis was performed using a time-lagged variable as a predictor. Let *W*_*t−d*_be the weekly aggregated average wastewater concentration at week in the aggregated Jefferson County, and represents a time lag.*H*_t_ represents the hospitalization rate at time *t*. The regression model with time lag dependent variable is given by:

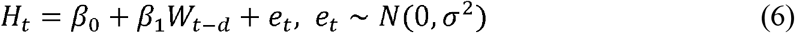

In this model, we changed the time lag d from 1 to 4 so that the maximum period from evidence of the community spread of COVID-19 in wastewater to reach a burden to hospitalization is about a month. Of note, hospitalizations data is available daily while wastewater is at a frequency of bi-weekly.

Additionally, we performed a simulation study using this regression model to check how much the hospitalization rate changes according to the vaccination rate. We changed the vaccination rate so that the vaccination percentage of the community was 0% and predicted the serial estimates Pred_t_ in Eq. (4). And then, we predicted the wastewater concentration using a linear regression model (5) and used that as the predictor in the regression model (6).

### S3.4. Calculation of effects based on factual and counterfactual scenarios

Effects of the factual and counterfactual (zero vaccinated or no Delta variant) are calculated using the area under the respective curves based on the models using factual (empirical) data and counterfactual (synthetic) data. The equation to estimate the effect is given as:

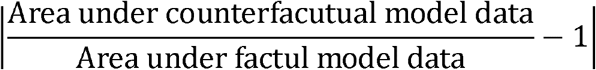

### S3.5. Sensitivity analysis for changing the amount of Delta variant transmission rate

In our analysis, we assumed the transmission rate of the Delta variant, denoted as β^, is 150% higher than the Alpha variant. Since this assumption is quite strong, to illustrate its effect, we conducted global sensitivity analysis under various alternative scenarios β * =γ β, Where β* and βare disease transmission rates in Eq. (1). We set > to 1.2, 1.5, 2.0, 2.5 and 3.0.Then we simulated the ODE (1) and calculate the basic reproduction number (*R*_0_).

### S3.6. The derivation of the basic reproduction number (R_0_)^6^

Using *SVI*_2_ *RT* model, let *X* be the vector of infected compartments, denoted by *x* =(*I* (1), *I* 2)T. The system has a disease-free state 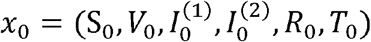. We define the matrix of new infection ℱ (*x*) and the matrix of all transitions except for the new infection *V*. The net transition rates are represented by *V(x)*.

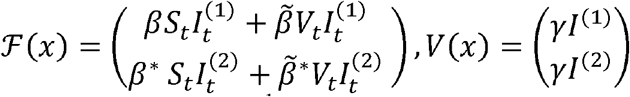

The next generation matrix is defined as FV^−1^ where F and V represent 2×2 matrices at *x*_0_ as follows:

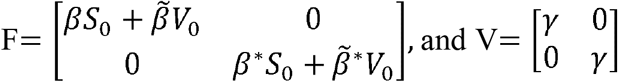

The next generation matrix K is calculated as

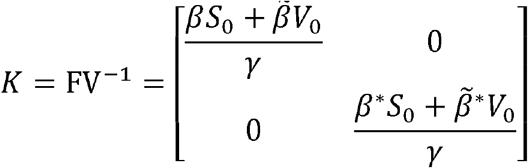

Finally, the basic reproduction number *R*_0_ is the maximum eigenvalue of the spectral decomposition of the next generation matrix K:

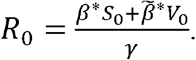

If we set *S*_0 =_ 1 and *V*_0_ = 0, then 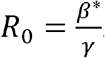.

## Appendix D. Wastewater variant detection

**Table S13.** Periods of Alpha and Delta variant wastewater dominance in sewersheds of Jefferson County, KY (USA). Dates determined by shift in major variant based on sampling schedule of wastewater collection.

| Sewershed | Alpha dominant in wastewater |  | Delta dominant in wastewater |  |
| --- | --- | --- | --- | --- |
|  | Start date | End date | Start date | End date |
| MSD1 | 3/30/21 | 5/17/21 | 7/12/21 | 8/30/21 |
| MSD2 | 3/30/21 | 5/24/21 | 7/12/21 | 8/30/21 |
| MSD3 | 3/30/21 | 6/21/21 | 7/19/21 | 8/30/21 |
| MSD4 | 3/30/21 | 7/5/21 | 7/19/21 | 8/30/21 |
| MSD5 | 3/30/21 | 6/28/21 | 7/26/21 | 8/30/21 |

